# Automated detection of axonal damage along white matter tracts in acute severe traumatic brain injury

**DOI:** 10.1101/2022.03.09.22271989

**Authors:** Chiara Maffei, Natalie Gilmore, Samuel B. Snider, Andrea S. Foulkes, Yelena G. Bodien, Anastasia Yendiki, Brian L. Edlow

## Abstract

New techniques for individualized assessment of white matter integrity are needed to detect traumatic axonal injury (TAI) and predict outcomes in critically ill patients with acute severe traumatic brain injury (TBI). Diffusion MRI tractography has the potential to quantify white matter microstructure *in vivo* and has been used to characterize tract-specific changes following TBI. However, tractography is not routinely used in the clinical setting to assess the extent of TAI, in part because focal lesions reduce the robustness of automated methods. Here, we propose a pipeline that combines automated tractography reconstructions of 40 white matter tracts with multivariate analysis of along-tract diffusion metrics to assess the presence of TAI in individual patients with acute severe TBI. We used the Mahalanobis distance to identify abnormal white matter tracts in each of 18 patients with acute severe TBI as compared to 33 healthy subjects. In all patients for which a FreeSurfer anatomical segmentation could be obtained (17 of 18 patients), including 13 with focal lesions, the automated pipeline successfully reconstructed a mean of 37.5 +/- 2.1 white matter tracts without the need for manual intervention. A mean of 2.5 +/- 2.1 tracts resulted in partial or failed reconstructions and needed to be reinitialized upon visual inspection. The pipeline detected at least one abnormal tract in all patients (mean: 9.07 +/- 7.91) and could accurately discriminate between patients and controls (AUC: 0.91). The individual patients’ profiles showed the number and neuroanatomic location of abnormal tracts varied across patients and levels of consciousness. The premotor, temporal, and parietal sections of the corpus callosum were the most commonly damaged tracts (in 10, 9, and 8 patients respectively), consistent with histological studies of TAI. TAI measures were not associated with concurrent behavioral measures of consciousness. In summary, we provide proof-of-principle evidence that an automated tractography pipeline has translational potential to detect and quantify TAI in individual patients with acute severe traumatic brain injury.

## 1. Introduction

Traumatic axonal injury (TAI) is the most common pathologic substrate of head trauma (Brody et al., 2015; McGinn & Povlishock, 2016) and has a strong association with adverse clinical outcomes (Benson et al., 2007; Gennarelli et al., 1982; Moe et al., 2020; Newcombe et al., 2010). TAI is caused by high-velocity translational and rotational forces that stretch and shear axons, leading to either primary axotomy or a secondary cascade that can result in further axonal injury (Hill et al., 2016). The diagnosis of TAI currently relies upon the neurological examination and the presence of visible white matter (WM) damage using routine neuroimaging techniques. However, due to its diffuse and microscopic nature (Adams et al., 1989; Blumbergs et al., 1995; Johnson et al., 2013), TAI is challenging to quantify using CT and conventional MRI (Betz et al., 2012), and its extent is likely underestimated in clinical practice. Furthermore, individual scores obtained from the neurological examination (*e*.*g*., the Glasgow Coma Scale) have limited diagnostic utility, given that neurologic deficits may reflect a broad spectrum of pathophysiologic processes.

For clinicians and the families of patients with acute severe traumatic brain injury (TBI), the lack of reliable diagnostic tools that measure the burden of TAI creates prognostic uncertainty and complicates discussions about continuation of life-sustaining therapy in the intensive care unit (ICU) (Izzy et al., 2013; Turgeon et al., 2011). Given the multifocal and heterogenous nature of TAI, neuroimaging studies that compare TBI patients at the group level are insensitive to TAI variability and therefore are suboptimal for gaining insight into TAI mechanisms in individual patients (Betz et al., 2012). New techniques for the individualized assessment of WM injury (Jolly et al., 2020; Kim et al., 2013; Yuh et al., 2014) are therefore needed to enhance detection of TAI and improve the accuracy of outcome prediction in critically ill patients with acute severe TBI.

Diffusion MRI (dMRI) probes WM tissue microstructure non-invasively (Basser et al., 1994), making it possible to identify TAI *in vivo* (Hashim et al., 2017; Li et al., 2011; Xu et al., 2007). Numerous studies have used dMRI to assess WM integrity in patients with TBI (Arfanakis et al., 2002; Asken et al., 2017; Galanaud et al., 2012; Hulkower et al., 2013; Inglese et al., 2005; Newcombe et al., 2007; Zhang et al., 2017) and have shown its sensitivity to detect WM abnormalities across TBI severities (*i*.*e*., mild, moderate, severe) and recovery phases (*i*.*e*., acute, subacute, chronic). Typically, summary metrics such as fractional anisotropy (FA) and mean diffusivity (MD) are extracted from WM regions of interest (ROIs) that are manually drawn in subject’s space (Inglese et al., 2005; Christine L. Mac Donald et al., 2011) or segmented from an atlas (Galanaud et al., 2012; O’Phelan et al., 2018). Whole-brain voxel-based analysis (Van Hecke et al., 2016) and tract-based spatial statistics (Smith et al., 2006) approaches are also used to compare normalized scalar maps between patient groups (Kinnunen et al., 2011), or between a control group and a patient (Jolly et al., 2020). While these approaches have demonstrated sensitivity to TAI (Jolly et al., 2020; Perlbarg et al., 2009), they do not assess injury of specific WM tracts at the individual level.

Diffusion-based tractography allows for the delineation and quantification of specific WM pathways at the individual level. As different WM tracts subserve different cognitive functions and their anatomical location varies across individuals, tractography methods may provide a more specific evaluation of WM damage that can inform early decisions about goals of care, continuation of life-sustaining therapy, and functional recovery. However, few studies have utilized tractography in patients with acute TBI (D’Souza et al., 2015; Ordóñez-Rubiano et al., 2017; Snider et al., 2019; Wang et al., 2008; Warner et al., 2010) or acute severe TBI (Ordóñez-Rubiano et al., 2017; Snider et al., 2019; Wang et al., 2008). Furthermore, dMRI tractography techniques are not routinely used in the clinical setting to assess the extent of TAI (Schweitzer et al., 2019) because the presence of focal lesions, the lower quality of the dMRI data compared to research settings, and the time feasibility constraints all limit the application of available tractography pipelines.

To address these barriers to clinical translation, we tested a pipeline for individualized TAI assessment in patients with acute severe TBI that combines automated tractography with a multivariate analysis of along-tract diffusion metrics. We used TRACULA (TRActs Constrained by UnderLying Anatomy), a method for global probabilistic tractography with anatomical neighborhood priors (Yendiki et al., 2011), to automatically reconstruct 40 WM tracts at the individual level (Maffei et al., 2021). The anatomical neighborhood priors in TRACULA encode information about the relative position of the tracts with respect to their surrounding anatomical structures, rather than their absolute coordinates in an atlas space. Therefore, TRACULA does not necessitate accurate registration to an atlas (Zollei et al, 2019; Maffei et al., 2021), which can be challenging in the presence of lesions and may not reflect interindividual anatomical differences. Furthermore, in contrast to local tractography methods that consider the local diffusion orientation at each step, TRACULA uses a global approach and would thus not “stop” in the presence of a lesion. We previously showed that TRACULA, when trained on high-quality data from the Human Connectome Project (HCP) (Fan et al., 2015), improves the accuracy of tractography reconstructions in routine-quality data when compared to a common multi-ROI tractography approach (Maffei et al., 2021). These methodologic advantages of TRACULA suggest potential for translation of this technique to critically ill patients with acute severe TBI, who are not medically stable enough to be scanned with a time-intensive, high-quality dMRI sequence.

We applied TRACULA to a dMRI dataset prospectively acquired in a cohort of 18 critically ill patients with acute severe TBI who were imaged on a clinical 3 Tesla MRI scanner. We used a multivariate approach based on the Mahalanobis distance to identify the abnormal WM tracts in each patient compared to 33 healthy subjects. The goals of this study were: i) to demonstrate the feasibility of applying the TRACULA automated tractography pipeline using dMRI data acquired on a clinical scanner in patients with acute severe TBI, including those with focal lesions; ii) to implement an individualized multivariate approach to measure patient-specific TAI severity, using reconstructions derived from the automated TRACULA tractography pipeline; and iii) to test for associations between bundle-specific dMRI measures of TAI and behavioral measures of consciousness.

## 2. Methods

### 2.1 Participants

We prospectively enrolled 18 patients (mean +/- standard deviation age: 28.6 +/- 8.7 years, 13 male) with acute severe TBI, sixteen of whom were enrolled as part of a previously described pilot study (Edlow et al., 2017), and two of whom were subsequently enrolled as pilot cases for an ongoing observational study (ClinicalTrials.gov NCT03504709; see Table 2 for additional information). Inclusion criteria were: 1) age 18 to 65 years; and 2) traumatic coma, defined by Glasgow Coma Scale (GCS) (Teasdale & Jennett, 1974) total score of six without eye opening on at least one neurologic examination before ICU admission; and 3) no eye opening for 24 hours after injury. Exclusion criteria were: 1) prior history of severe brain injury or neurodegenerative disease; 2) life expectancy less than six months per physician judgment; 3) presence of metal contraindicating MRI; and/or 4) no pre-injury English fluency.

**Table 2.** Demographics and Clinical Characteristics of the TBI patients. CRS-R: Coma Recovery Scale-Revised; CRS-R-T: Total score of the CRS-R; CRS-R-M: Motor CRS-R sub-scale; CRS-R-A: Auditory CRS-R sub-scale; CRS-R-O/V: Oromotor-verbal CRS-R sub-scale; GCS: Glasgow Coma Scale; GCS-T: Total score of the GSC; LOC: level of consciousness; MCS+ = minimally conscious state plus, evidence of language function (*e.g*., reproducible movement to command, object recognition, intelligible verbalization); MCS- = minimally conscious state minus, no evidence of language function, but shows at least one behavior consistent with a minimally conscious state (*e.g*., automatic motor response); MVA: motor vehicle accident; Ped vs car: pedestrian versus car; PTCS: post-traumatic confusional state; SD: standard deviation; VS: vegetative state. *Unable to determine exact date based on intensive care unit clinician assessments. We used 1 day for further analysis. **Patients who died in the ICU after withdrawal of life sustaining therapy and before command-following was observed.

| ID | Age Range | Sex | Days in Coma Range | Days Until Command - Following | Days to MRI Range | LOC at MRI | GCS-T at MRI | CRS-R-T at MRI | CRS-R-M at MRI | CRS-R-A at MRI | CRS-R-O/V at MRI |
| --- | --- | --- | --- | --- | --- | --- | --- | --- | --- | --- | --- |
| P1 | 26-30 | M | 1-5 | 1 | 16-20 | PTCS | 15 | 23 | 6 | 4 | 3 |
| P2 | 21-25 | M | 1-5 | 6 | 1-5 | MCS- | 7 | 4 | 3 | 0 | 1 |
| P3 | 16-20 | F | 1-5 | 10 | 1-5 | Coma | 5 | 1 | 1 | 0 | 0 |
| P4 | 16-20 | M | 1-5 | 2 | 16-20 | PTCS | 14 | 23 | 6 | 4 | 3 |
| P5 | 31-35 | M | 6-10 | 26 | 11-15 | VS | 7 | 3 | 0 | 0 | 2 |
| P6 | 26-30 | F | 1-5 | 9 | 6-10 | VS | 9 | 6 | 2 | 0 | 1 |
| P7 | 41-45 | M | 1-5 | 6 | 11-15 | MCS+ | 13 | 18 | 5 | 3 | 3 |
| P8 | 31-35 | M | 1-5 | 7 | 6-10 | PTCS | 11 | 20 | 5 | 4 | 2 |
| P9 | 31-35 | M | 1-5 | 8 | 11-15 | MCS+ | 10 | 9 | 2 | 3 | 1 |
| P10 | 21-25 | M | 1-5 | 7 | 11-15 | MCS- | 10 | 10 | 5 | 1 | 1 |
| P11 | 21-25 | F | 1-5 | 9 | 11-15 | PTCS | 14 | 22 | 6 | 4 | 3 |
| P12 | 26-30 | F | 11-15 | ** | 6-10 | Coma | 5 | 1 | 1 | 0 | 0 |
| P13 | 16-20 | M | 1-5 | 2 | 1-5 | MCS+ | 10 | 12 | 5 | 3 | 1 |
| P14 | 51-55 | M | 6-10 | ** | 6-10 | VS | 6 | 3 | 1 | 0 | 0 |
| P15 | 26-30 | M | 1-5 | 8 | 6-10 | MCS- | 7 | 3 | 3 | 0 | 0 |
| P16 | 31-35 | M | 1-5 | 3 | 1-5 | MCS+ | 10 | 12 | 5 | 4 | 0 |
| P17 | 26-30 | F | 1-5 | ** | 11-15 | VS | 8 | 3 | 1 | 0 | 0 |
| P18 | 26-30 | M | 1-5 | 0-2* | 26-30 | PTCS | 11 | 18 | 6 | 3 | 2 |
| **Mean (SD)** | 28.56 (8.71) |  | 3.00 (3.25) | 7.0 (6.1) | 10.39 (6.46) |  | 9.6 (3.1) | 10.6 (8.1) | 3.5 (2.2) | 1.8 (1.8) | 1.3 (1.2) |
| **Range** | 18-51 |  | 1-13 | 1-26 | 1-28 |  | 5-15 | 1-23 | 0-6 | 0-4 | 0-3 |

We also enrolled 33 healthy control subjects (mean +/- standard deviation age 30.93 +/- 8.75 years, 21 male) with no history of neurological, psychiatric, cardiovascular, pulmonary, renal or endocrinological disease (Edlow et al., 2017). We confirmed that age was not statistically different between the two groups using a nonparametric Wilcoxon sum rank test in Python 3 using SciPy Stats 1.3.1 (Jones et al., 2001) (*W* = −1.116, *p* = 0.26). Written informed consent was obtained from healthy subjects and from patients’ surrogate decision-makers in accordance with a study protocol approved by the Mass General Brigham Institutional Review Board.

### 2.2 Behavioral Assessment of Consciousness

Immediately prior to MRI, each patient’s level of consciousness (LOC) was prospectively assessed by a neurologist on the investigator team (B.L.E.) via behavioral evaluation with the GCS (Teasdale & Jennett, 1974) the Coma Recovery Scale-Revised (CRS-R) (Giacino et al., 2004), and the Confusion Assessment Protocol (CAP) (Sherer et al., 2005). Per routine ICU care at our institution, neurological examinations and GCS assessments were performed off of sedation every two-to-four hours (depending on clinical stability and safety considerations) by treating clinicians. ICU clinician GCS assessments were used to determine time from injury to coma emergence (*i.e*., eye opening or localization to noxious stimulation) and time to command-following. If a patient’s neurological examination fluctuated, the first behavioral evidence of coma emergence and command-following were reported. The GCS Total was included as a primary dependent variable in this study given its widespread use clinically; however, its limitations (i.e., only moderate psychometric properties, administration process is not standardized) must be acknowledged (Bodien et al., 2021). The CRS-R, which has strong psychometric properties and a standardized administration process (Giacino et al., 2004; Teasdale et al., 2014), along with the CAP, were used in addition to the GCS to more precisely classify each patient’s LOC into the following categories: coma, vegetative state, minimally conscious state minus (MCS-; MCS without language function), MCS plus (MCS+; MCS with language function), and post-traumatic confusional state (PTCS).

### 2.3 MRI Data Acquisition

MRI data were acquired as soon as the treating clinicians deemed patients stable for transport. Diffusion-weighted imaging data were acquired in the ICU on a 3T Skyra scanner (Siemens Medical Solutions, Malvern, PA) with a 32-channel head coil. Three healthy subjects and eight patients were scanned using an echo-planar imaging (EPI) sequence with the following parameters: 2×2×2 *mm*, 60 *b* = 2,000 *s* / *mm*^2^ and 10 *b* = 0 *s* / *mm*^2^ volumes, *TR* = 13,700 ms, and *TE* = 98 *ms*. Thirty healthy subjects and 8 patients were scanned using an EPI sequence with simultaneous multi-slice (SMS) acceleration (Setsompop et al., 2012) and the following parameters: 2×2×2 *mm*, 60 *b* = 2000 *s*/*mm*^2^ and 10 *b* = 0 *s* / *mm*^2^, *TR* = 6,700 *ms, TE* = 100 *ms*, acceleration factor = 2. Previous work from our group showed that diffusion-derived connectivity metrics did not differ significantly between these two acquisitions (Snider et al., 2019). The length of the diffusion sequence was 8 minutes and 46 seconds. Multi-echo MPRAGE structural images (van der Kouwe et al., 2008) were also acquired with the following parameters: 1×1×1 *mm*, acquisition matrix = 256 × 256, *TE* = 1.69 *ms, TR* 2530 *ms, flip angle* = 7°. All patients and healthy subjects were imaged using the same scanner and head coil.

### 2.4 Diffusion MRI Data Processing

Diffusion weighted images were skull stripped and corrected for eddy-current distortions and movement in FSL 6.0.1 (Andersson & Sotiropoulos, 2015). The tensor and the ball-and-stick model were fit to the data using DTIFIT and BEDPOSTX (Behrens et al., 2003) in FSL 6.0.1, respectively. Automated reconstruction of 40 major WM pathways was performed using the global probabilistic tractography algorithm TRACULA in FreeSurfer 7.2.0 (Maffei et al., 2021; Yendiki et al., 2011). Although TRACULA can reconstruct a total of 42 WM tracts, we decided to exclude the right and left fornix, because optimal identification of these WM bundles requires segmentation of thalamic subnuclei that are not provided by the standard “recon-all” FreeSurfer processing pipeline. The mathematical formulation of TRACULA has been described elsewhere (Yendiki et al., 2011, 2016). Briefly, the algorithm models a pathway as a cubic spline, which is initialized with the median streamline of the training set. A random sampling algorithm is used to draw samples from the posterior probability distribution of the pathway, which is decomposed into the likelihood term and the prior term. The likelihood term fits the shape of the spline to the diffusion orientations obtained from the ball-and-stick model in the voxels that it goes through. The prior term fits the shape of the spline to its anatomical neighborhood, given the manually labeled examples of this pathway from the training set and the anatomical segmentation volumes of both test and training subjects. The number of control points of the cubic spline were chosen as previously described (Maffei et al., 2021).

Along-tract tensor-derived metrics obtained from DTIFIT were extracted for each of the reconstructed tracts to perform a pointwise assessment of streamline tractography attributes (Jones et al., 2005). For each of the 40 tracts, 1D along-tract profiles of FA and MD were generated by projecting the value of each measure from every point on every automatically reconstructed streamline to its nearest point on a reference streamline, as previously described (Maffei et al., 2021). The reference streamline is the mean of the training streamlines for each tract, ensuring that all subject data were sampled at the same number of cross-sections along a given bundle. Before extracting the diffusion measures, the posterior probability distribution estimated by TRACULA was thresholded by masking out all values below 20% of the maximum, which is the default threshold in TRACULA (Maffei et al., 2021; Yendiki et al., 2011; Zöllei et al., 2019). All pre-processing steps detailed above are performed automatically by the TRACULA pipeline.

### 2.5 Structural MRI Data Processing

The anatomical segmentation that was needed to compute the anatomical neighborhood priors in TRACULA was obtained by analyzing the structural T1-weighted image in FreeSurfer 7.1.1 (Dale et al., 1999; Fischl, 2012). The automated “recon-all” pipeline was run with default parameters, except for the “bigventricle” option, which was added to optimize segmentation in patients that may have enlarged ventricles post-injury. The obtained segmentation volumes included a combination of the Desikan-Killiany cortical parcellation labels (Desikan et al., 2006) and the standard FreeSurfer subcortical segmentation. To compute the prior probabilities on the anatomical neighbors of the tracts using TRACULA, the anatomical segmentations were transformed to the subject’s individual dMRI space. This within-subject, dMRI-to-T1 alignment was performed using a boundary-based, affine registration method (Greve & Fischl, 2009). To ensure that the relative position of the anatomical structures was the same for all subjects, and to map the median streamline from the training data to the subject during initialization, all subjects’ images were mapped onto a template brain. We used the non-linear symmetric normalization (SyN) in ANTs (Avants et al., 2008) to map each subject’s images onto an FA template constructed from the training dataset (Maffei et al., 2021). It is important to highlight that this step is only used to initialize the reconstruction of the tract, which is then refined by fitting it to the anatomy of the individual subject.

### 2.6 Quality assessment of tract reconstructions

To assess the feasibility of implementing TRACULA in a clinical setting, we performed a qualitative assessment to evaluate the accuracy of the tract reconstructions in patients with acute severe TBI. We considered the application of TRACULA in this population to be feasible if more than 50% of the automated reconstructions were successful. For this qualitative assessment, a study investigator (C.M.) visually inspected all 40 tracts for all patients. Reconstructions were considered successful if the tracts traversed the WM regions and reached the cortical areas used to define these tracts in the manual labeling protocols. These protocols were carefully defined by the known anatomy of each tract and are detailed in (Maffei et al., 2021).

When a focal lesion was present, the reconstruction was considered successful if the tract reached the cortical termination regions delineated in (Maffei et al., 2021), even if the tract’s course was altered by the presence of the lesion. As TRACULA uses a global probabilistic approach to estimate the tracts, post-processing steps to remove false positive reconstructions were not necessary. However, when a suitable solution for the initial reconstruction of the pathway is not found, the tract appears as a single curve (See Figure 1 for a simulated example). This can be due to errors in the subject-to-template registration step, or to a misplacement of the FreeSurfer segmentation labels in the presence of lesions. We refer to these reconstructions as “failed reconstructions”. Moreover, as a single threshold is used across all tracts and groups to threshold the posterior probability of the tract, some reconstructions can result in incomplete tracts or in unusually small tracts for which many voxels did not survive the threshold (See Figure 1 for an example). We refer to these reconstructions as “partial reconstructions”.

**Figure 1.**
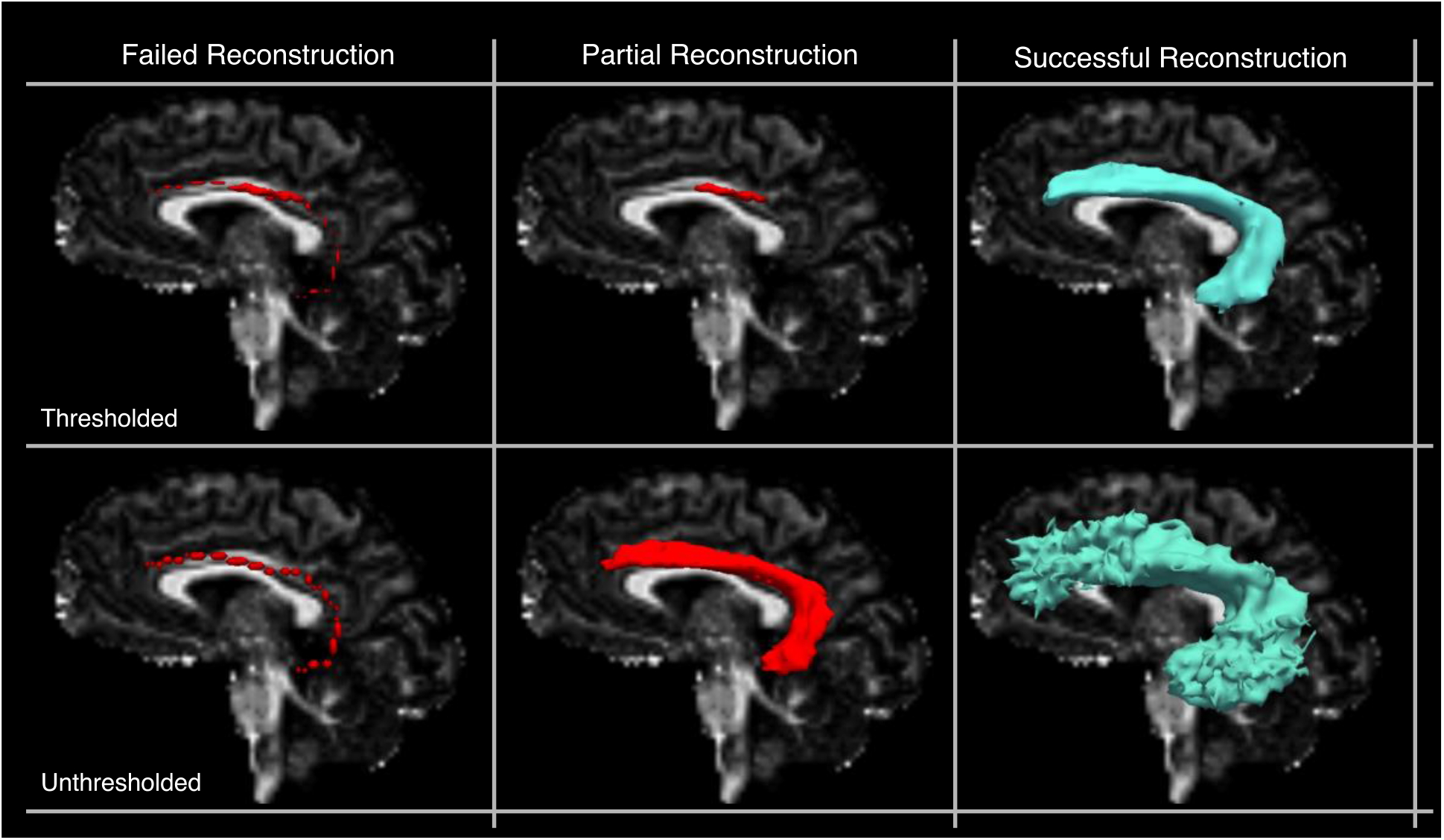
Simulated examples of failed, partial, and successful TRACULA reconstructions for a representative tract (the left arcuate fasciculus) in a control subject. Reconstructions are shown thresholded (20% of the maximum posterior probability) and unthresholded. Reconstructions are shown on sagittal fractional anisotropy scalar maps.

We reran TRACULA on the tracts that resulted in either failed or partial reconstructions by reinitializing the control points of the initial spline, and setting the *reinit* parameter to 1 (Yendiki et al., 2011). We retained the re-initialized reconstructions if they resulted in correct tracts as defined above. The same criteria were applied to patients and healthy subjects. This was the only manual intervention performed in the TRACULA pipeline. Tracts that had partial or failed reconstructions after reinitialization were excluded from subsequent analyses.

### 2.7 Multivariate analysis

Figure 2 shows the steps of the multivariate analysis. To measure the extent of TAI in each patient, we employed a multivariate analysis that included along-tract measures of both FA and MD. We focused on along-tract measures of FA and MD, as these microstructural measures are the most commonly studied in the field of TBI (Hulkower et al., 2013; Mac Donald et al., 2007). For this analysis we used both the tracts that had been successfully reconstructed without need of reinitialization and the tracts that needed to be reinitialized (*Section 2.6*). For each subject, each tract was divided uniformly in *i* = 1,.., 4 segments and the along-tract measures of FA and MD, extracted at each point along the tract in TRACULA (see *Section 2.4*), were averaged within each segment to obtain a total of eight features for each tract (*FA*_1_, *FA*_2_, *FA*_3_, *FA*_4_, *MD*_1_, *MD*_2_, *MD*_3_, *MD*_4_). We collapsed the along-tract measurements in four equidistant segments to ensure that, for each tract, the number of features (*i.e*., the measures for each tract; *m* = 8) was smaller than the number of observations (*i.e*., the number of healthy control subjects; *n* = 33). To meet normality assumptions, we conducted Shapiro-Wilk tests in Python 3 using SciPy Stats 1.3.1 (Jones et al., 2001) and tested for the normality of *FA*_*i*_ and *MD*_*i*_ for each of the 40 tracts in the control group. For the tracts for which the distribution of *FA*_*i*_ or *MD*_*i*_ departed significantly from normality (*p* < 0.05) we applied a rank-based inverse normal transformation to the data for both healthy controls and patients in Python 3 (Blom constant = 3 / 8, (Blom, 1958)), as in (Jolly et al., 2020).

**Figure 2.**
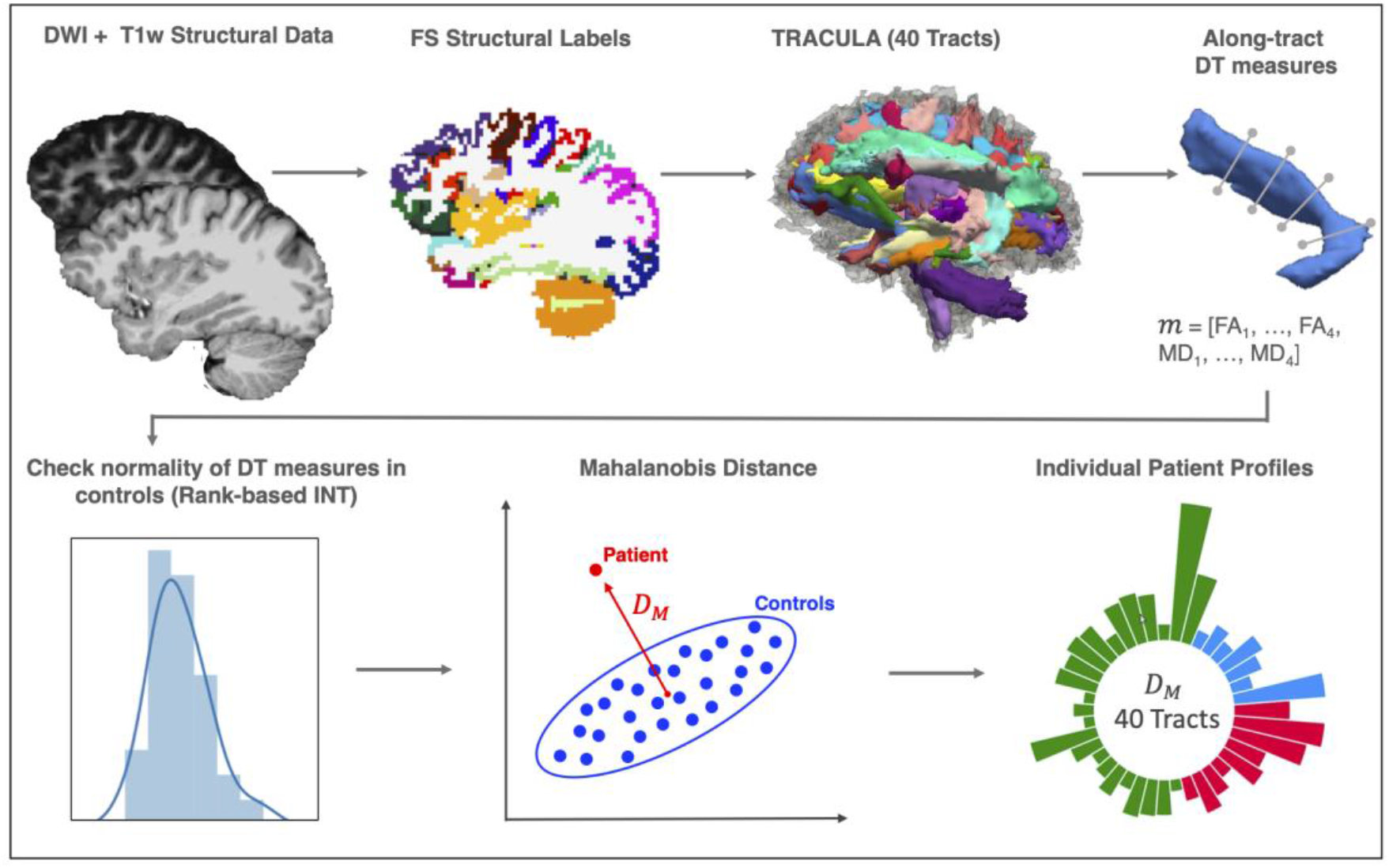
Schematic of the automated individualized pipeline. After pre-processing of diffusion weighted imaging (DWI) and structural T1-weighted imaging data, which included extraction of the FreeSurfer (FS) anatomical labels, the 40 white matter tracts were reconstructed in TRACULA. Along-tract measures derived from the diffusion tensor (DT) were then extracted for four equidistant segments for each of the 40 tracts. We checked the normality of the distribution for these measures in the controls and applied a Rank-based inverse normal transformation (INT) to the tracts for which assumptions of normality were not met (*p* < 0.05). We then computed the Mahalanobis distance (*D*_*M*_) for each tract using along-tract fractional anisotropy (FA) (*FA*_1_, …, *FA*_4_) and mean diffusivity (MD) (*MD*_1_,…, *MD*_4_) as features (*m* = 8) (here shown as a simplified 2D example). Finally, we built individual patient profiles based on the distance of each patient from the controls for each of the 40 white matter tracts.

For each of the 40 WM tracts, we compared the 8-D feature vector of dMRI measures (*FA*_1,…,4_ and *MD*_1,…,4_) between each patient and the healthy population based on the Mahalanobis distance *D*_*M*_, which is defined as (Mahalanobis, 1936):

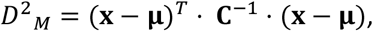

where **x** is the feature vector of a specific WM tract in a patient, **μ** is the mean feature vector of the same tract over controls, and **C** is the *m* × *m* covariance matrix of the feature vector in controls. The values of *D*^2^_*M*_ reflect how far a patient, represented as a point in the 8-D feature space, is from the normative distribution, which is estimated from the healthy controls. The Mahalanobis distance has been used in neuroimaging studies, *e.g*., to classify between patients with neurological diseases and controls (Lindemer et al., 2015) and to detect individual neurodevelopmental differences (Dean et al., 2017). We computed the *D*_*M*_ between each individual patient and the healthy controls for each of the 40 tracts to build individual profiles and identify the location and severity of WM abnormalities in patients. For multivariate Gaussian data, the distribution of *D*^2^_*M*_ values is known to be Chi-squared with degrees of freedom equal to the number of features (Gnanadesikan & Kettenring, 1972). We computed the p-values corresponding to the Chi-square statistic in Python 3 using SciPy Stats 1.3.1 (Jones et al., 2001) and considered a WM tract to be abnormal if its *p* − *value* was < 0.001 (*p* < 0.05, Bonferroni adjusted for multiple comparisons across 40 tracts). We extracted the total number of abnormal tracts for each patient.

To measure the accuracy of the pipeline in the task of detecting TAI, we computed its performance in discriminating between patients and controls. First, we computed the *D*_*M*_ between each control and the remaining control population in a leave-one-out fashion. For each of the 33 health controls, we used the multivariate distribution of along-tract dMRI measures (*FA*_1,…,4_ and *MD*_1,…,4_) from the remaining 32 heathy controls to compute the *D*_*M*_ of each WM tract and extract the total number of abnormal tracts at *a* = 0.001. We used a Wilcoxon rank sum test in Python 3 (SciPy Stats 1.3.1) to assess differences in the number of abnormal tracts between patients and controls. We then performed a receiver operating characteristic (ROC) analysis by varying simultaneously the alpha threshold used to obtain the number of abnormal tracts (*range* = 0.0001: 0.05, *step* = 0.001) for each subject, and the number of abnormal tracts used to label a subject as control or patient (*range* = 1: 40, *step* = 1). For each combination of alpha and number of abnormal tracts, we computed the false positives (FP; number of controls classified as patients), the false negative (FN; number of patients classified as controls), the true positives (TP; number of patients classified as patients), and the true negatives (TN; number of controls classified as controls) to compute the true-positive rate (*TPR; TP*/(*TP* + *FN*)) and false-positive rate (*FPR*; *FP*/ (*FP* + *TN*)). We obtained the ROC curve by plotting the TPR as a function of FPR and computed the area under the ROC curve (AUC) to quantify accuracy. To visualize the difference in distribution of *D*^2^_*M*_ values between the two populations we plotted the probability density functions of *D*^2^_*M*_ values for patients and controls for each of the 40 tracts.

Finally, we compared the results obtained using all tracts – those that were successfully reconstructed and those that were reinitialized – to the results obtained using only the tracts that were successfully reconstructed without reinitialization (Section 2.6). We used a Wilcoxon signed-rank test in Python 3 (SciPy Stats 1.3.1) to assess differences in the number of abnormal tracts and their mean *D*_*M*_ per patient.

### 2.8 Testing for the effect of head motion

To ensure differences in *D*^2^_*M*_ values between patients and controls were not influenced by head motion in the scanner, we first computed a total motion index (TMI) for each subject as described in (Yendiki et al., 2014). The TMI is a composite score of four motion measures: i) average volume-by-volume translation, ii) average volume-by-volume rotation, iii) percentage of slices with signal drop-out, and iv) signal drop-out severity. For each subject, the TMI was computed in the following manner:

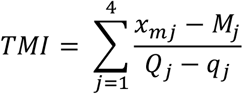

where *j* = 1, …, 4 are the four motion measures listed above, *x*_*mj*_ is the value of the *j*-th motion measure for the *m*-th subject and *M*_*j*_, *Q*_*j*_, and *q*_*j*_ are respectively the median, upper quartile, and lower quartile of the *j*-th measure over all the subjects (Yendiki et al., 2014). We then used a nonparametric Wilcoxon rank sum test in Python 3 (SciPy Stats 1.3.1) to assess if there were statistically significant differences in head motion between the patients and controls. Finally, we conducted a linear mixed effects regression model in R (RStudio Team, 2020) using the ‘lmer’ function in the “lme4” package to test if TMI significantly predicted *D*^2^_*M*_, while controlling for group (i.e., control, patient) and tract (*i.e*., 40 reconstructed tracts), and including participant as a random intercept.

### 2.9 Testing the relationship with behavioral measures

First, we performed non-parametric Spearman correlations to support variable selection and model building for the primary regression analyses. A correlation matrix was constructed in R (RStudio Team, 2020) using the ‘cor’ function in the “stats” package. The matrix consisted of the dependent variables (*i.e*., GCS Total, CRS-R Total, days in coma as defined by the GCS, days to command-following as defined by the GCS), the independent variables (*i.e*., average D_M_ of affected tracts and number of affected tracts), and potential confounding variables (*i.e*., age, TMI, days to MRI) (Edlow et al., 2016). P-values for the correlations were not corrected for multiple comparisons; rather, the significance level was maintained at 0.05 to ensure the identification of any confounding variables.

Next, linear regressions were planned for completion in R using the ‘lm’ function (RStudio Team, 2020). Separate linear regression models would be conducted for each of the dependent variables (i.e., GGS Total, CRS-R Total, days in coma, days to command-following) with the average D_M_ of affected tracts and the number of affected tracts as independent variables, and any identified confounding variables as covariates. Predictor variables that were significantly correlated with one another would be included in separate models to avoid multicollinearity.

Finally, an ordinal logistic regression analysis was applied to predict LOC, given the categorical nature of this variable as defined by the CRS-R (*i.e*., coma, VS, MCS-, MCS+, PTCS). The ordinal logistic regression was performed in R (RStudio Team, 2020) using the ‘plor’ function from the “MASS” package (Venables & Ripley, 2002) with LOC as the dependent variable; the average D_M_ of affected tracts and the number of affected tracts as independent variables, and any identified confounding variables as covariates (*i.e*., determined via univariate ordinal logistic regression analyses between LOC and age, days to MRI, and TMI).

## 3. Results

### 3.1 Patient Demographics and Clinical Characteristics

Patient demographics and clinical characteristics are provided in Table 2, including clinical indicators of coma duration (*i.e*., days in coma, days until command-following, level of consciousness at MRI); and behavioral measures of consciousness (*i.e*., GCS and CRS-R total scores), including subscales measuring language and motor function.

### 3.2 Quality assessment and feasibility of tractography reconstructions

Figure 3 shows the reconstruction of the 40 tracts for two representative patients, one without focal lesions and one with a large right frontal hemorrhagic contusion. We observed that the TRACULA pipeline successfully reconstructed 93% of the tracts (mean: 37,5; SD: 2.18) across all patients without manual intervention, exceeding our feasibility threshold of 50% by 43% (Section 2.6).

**Figure 3.**
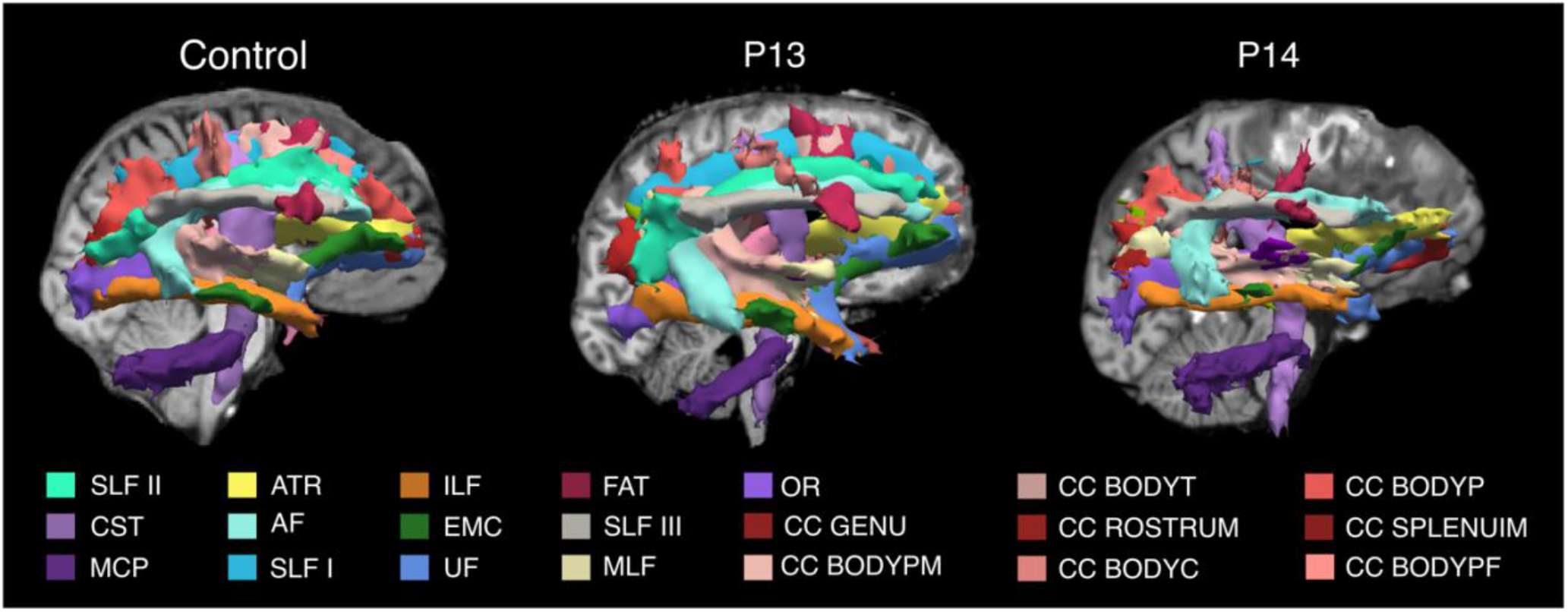
TRACULA reconstructions. The 40 tracts automatically reconstructed in TRACULA are shown in 3D sagittal view (right hemisphere) for one control and two patients, one with no focal lesions (P13), and one with a focal lesion in the right frontal lobe (P14). For each subject, tracts are overlaid on the T1-weighted structural volume. Color labels are reported only for tracts visible in the figure. **Complete list of WM tracts abbreviations:** ACOMM: anterior commissure; AF: arcuate fasciculus; AR: acoustic radiation; ATR: anterior thalamic radiation; CC BODYC: central section of the body of the CC; CC BODYP: parietal section of the body of the CC; CC BODYPF: prefrontal section of the body of the CC; CC BODYPM: premotor section of the body of the CC; CC BODYT: temporal section of the body of the CC ; CC GENU: genu of the CC; CC ROSTRUM: rostrum of the CC; CC SPLENIUM: splenium of the CC; CBD: dorsal portion of cingulum bundle; CBV: ventral portion of the cingulum bundle; CC: corpus callosum; CST: cortico-spinal tract; EMC: extreme capsule; FAT: frontal Aslant tract; ILF: inferior longitudinal fasciculus; LH: left hemisphere; ILF: middle longitudinal fasciculus; MVA: motor vehicle accident; OR: optic radiation; Ped vs car: pedestrian versus car; RH: right hemisphere; SLF I,II,III: first, second, and third branch of the superior longitudinal fasciculus; UF: uncinate fasciculus.

Figure 4 reports the number of reconstructions for each patient that i) were reconstructed without the need for reinitialization, ii) resulted in either partial or failed reconstruction and were reinitialized successfully, and iii) resulted in partial or failed reconstructions even after reinitialization (See *Section 2.6*). For a complete list of tracts that were reinitialized for each patient see supplementary table S1. In one patient (P1) the FreeSurfer “recon-all” stream failed due to the presence of a massive lesion that injured a substantial portion of the cerebral cortex in the right hemisphere. This patient was thus excluded from the TRACULA pipeline and is not reported in further results. For five of the remaining 17 patients (P2, P3, P6, P12, P16), reconstructions were complete and did not require reinitialization, even in the presence of focal lesions (P2, P6, P12) (Supplementary figure S1). For the remaining twelve patients, a small number of tracts (mean = 2.47, range = 1:7) resulted in failed or partial reconstructions and required reinitialization. For nine patients out of twelve, reinitialization led to successful reconstructions (as defined in Section 2.6). For one of these twelve patients (P4) two tracts were only partially reconstructed after reinitialization (MCP, LH CST), and for two patients (P10, P14) two and three tracts respectively (P10: ACOMM, RH UF; P14: ACOMM, RH SLFII, CC BODYPF) did not improve after reinitialization and resulted in failed reconstructions (Figure 3, P14). Upon visual inspection, failed or partial reconstructions were attributable to hemorrhagic contusions in the temporal and frontal lobes, respectively, which caused disruption of three tracts each (Supplementary figure S2).

**Figure 4.**
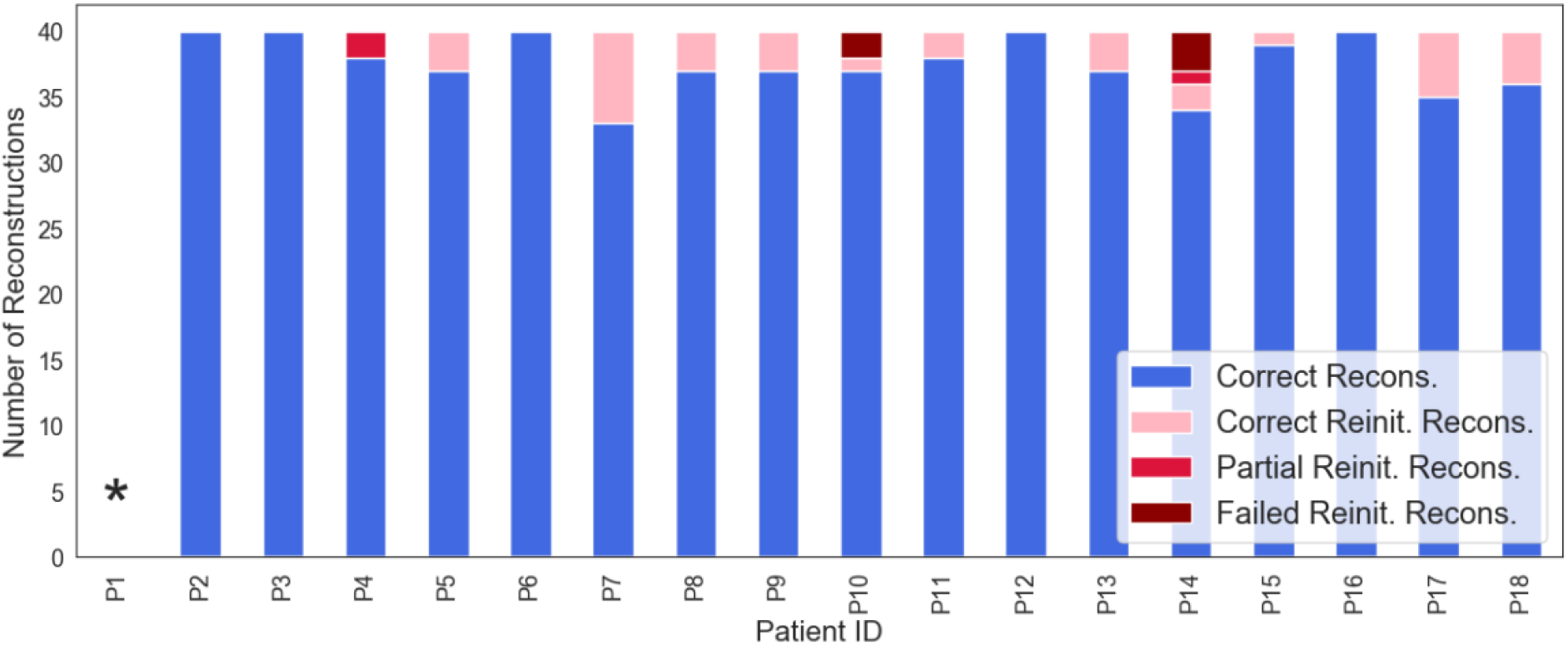
The number of correct reconstructions that did not need to be reinitialized (blue), the number of correct reconstructions after reinitialization (pink), and number of tracts that resulted in partial (red) or failed (maroon) reconstructions even after reinitialization are shown for each patient. * For P1, the FreeSurfer automated segmentation pipeline failed due to the presence of a massive lesion, which made it infeasible to run the TRACULA pipeline. Recons: reconstructions; Reinit: reinitialized.

### 3.3 Multivariate individualized assessment of white matter damage

The Shapiro-Wilk test showed that the distribution of *FA*_*i*_ and *MD*_*i*_ departed significantly from normality (p-value < 0.05) in 8.6 tracts per measure on average in the controls. Supplementary table 2 reports the tracts along with the *W* − and *p* − *value*. We applied a rank-based inverse normal transformation to these tracts for both healthy controls and patients before proceeding with the multivariate analysis (Section 2.7). Figure 5 shows the *D*^2^_*M*_-based individual profiles obtained from the multivariate analysis for four representative patients at different levels of consciousness (Coma, VS, MCS, PTCS). Individual profiles for all 17 patients are shown in Supplementary figure S3. The individual profiles show the Mahalanobis distance of each of the 40 WM tracts from the multivariate distribution of diffusion measures (along-tract FA and MD values) for that tract in the control population. We considered a WM tract to be abnormal if its distance exceeded the critical value of the Chi-squared statistic at *α* = 0.001 (pink dashed line in figure 5) (Section 2.7). The number and type of abnormal tracts varied across LOC (Figure 5, supplementary figure S3). To investigate whether a common pattern of TAI localization was visible across individuals, we grouped the reconstructed WM tracts as follows: i) commissural tracts: ACOMM, CC-BODYC, CC-BODYT, CC-BODYPF, CC-BODYP, CC-BODYPM, CC-GENU, CC-ROSTRUM, CC-SPLENIUM; ii) projection tracts: CST, ATR, AR, OR; iii) association tracts: all the other WM tracts. Commissural tracts were shown to have overall higher *D*^2^_*M*_ values (Supplementary figure S4) that resulted in a higher number of significantly extreme values (*p* < 0.001) across patients compared to projection and association tracts (Supplementary figure S5). Specifically, the CC-BODYPM, the CC-BODYP, and CC-BODYT resulted abnormal in 10, 9, and 8 patients respectively. For each patient we extracted the total number of abnormal tracts. On average, 9.07 +/- 7.9 SD tracts were abnormal in patients with TBI (Figure 6).

**Figure 5.**
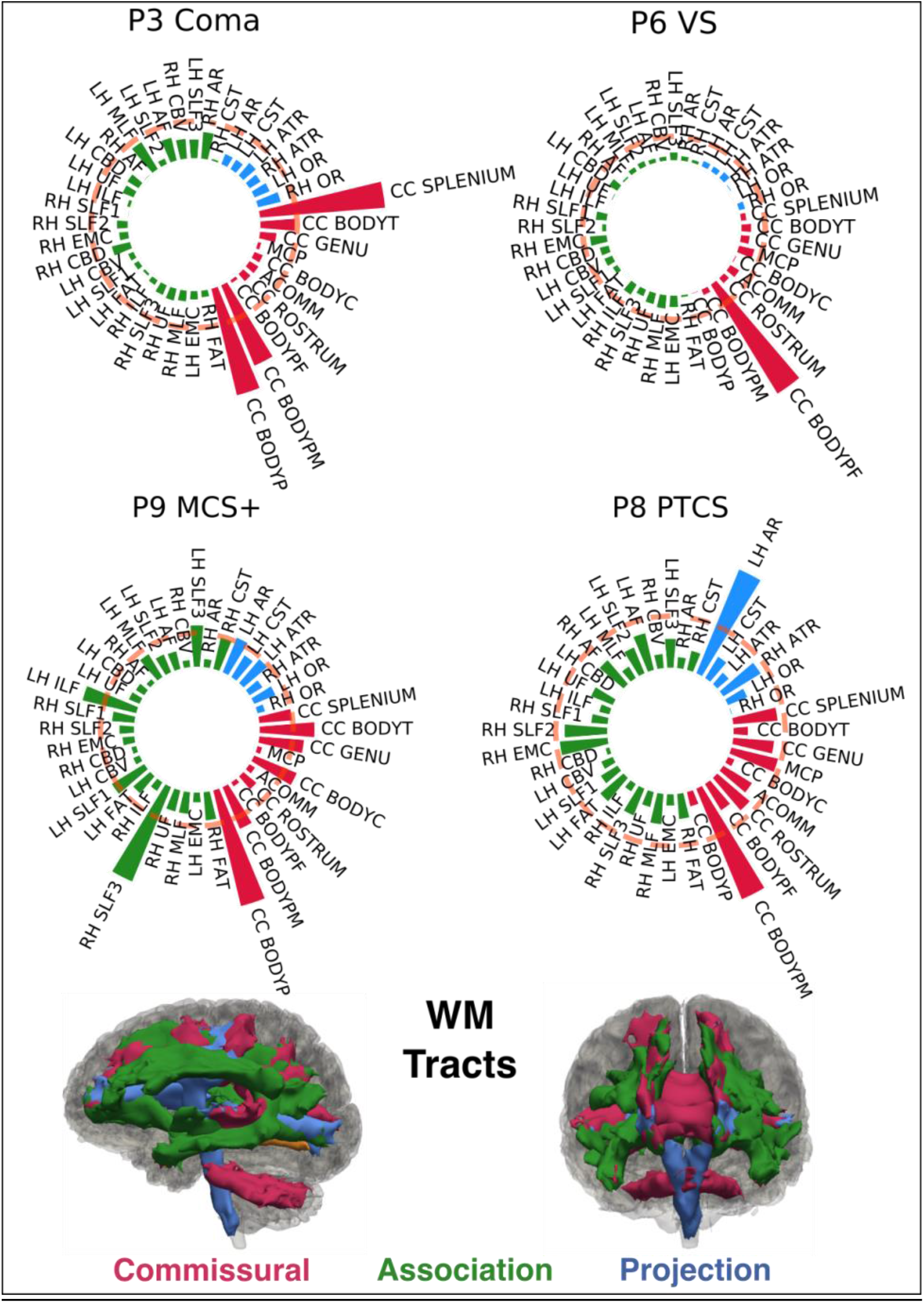
Individual Tract Profiles. The figure shows the individual profiles for four representative subjects, each with a different level of consciousness (Coma, VS, MCS+, PTCS). The Mahalanobis distance for the reconstructed tracts is plotted using polar bar plots. The pink dashed line corresponds to the critical value of the Chi-squared statistic at *α* = 0.001. Bars that extend beyond the dashed line represent tracts for which the multivariate diffusion measures (along-tract FA and MD values) were significantly different from the multivariate distribution of those measures in controls and were considered damaged. White matter tracts are grouped in commissural (red), associative (green), and projection pathways (blue). The 3D reconstructions of the 40 WM tracts are shown for a representative control subject in sagittal and coronal view, color coded based on the group they belong to. MCS+: minimally conscious state plus; PTCS: post-traumatic confusional state; VS: vegetative state; WM: white matter. For a complete list of WM tract abbreviations see Figure 3.

**Figure 6.**
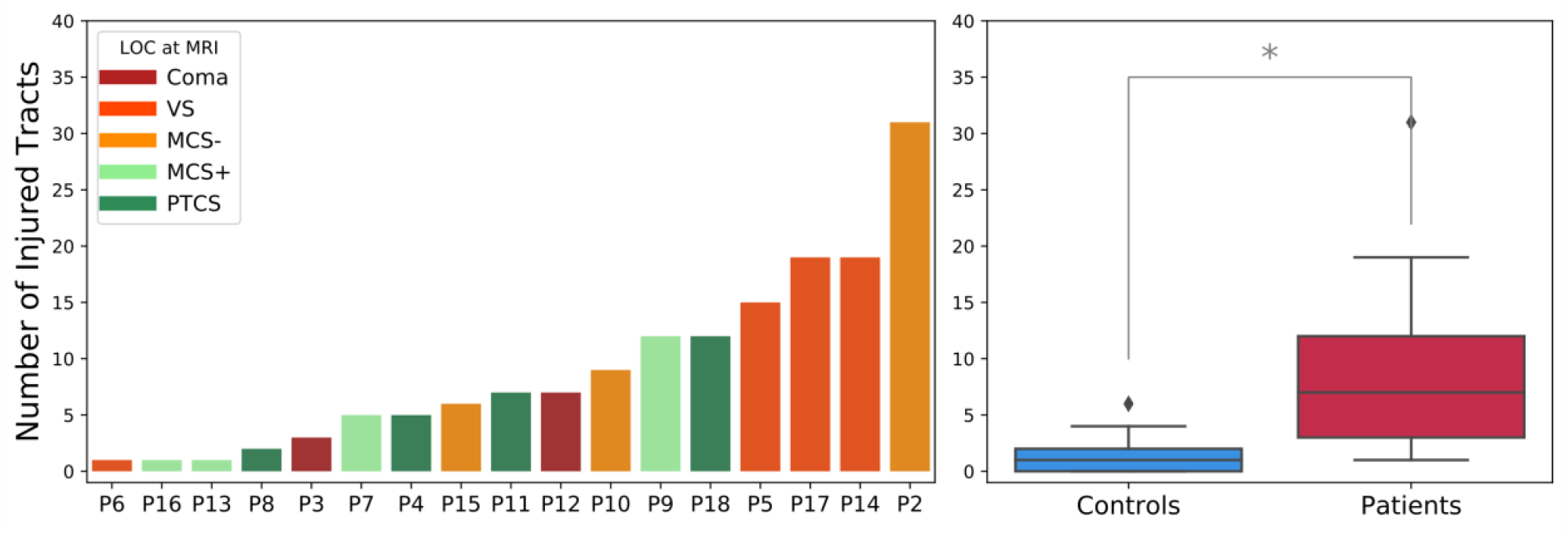
Number of affected tracts per subject. Left panel: The number of abnormal WM tracts is shown for each patient with TBI. A tract was considered abnormal if its *p* − *value*, corresponding to a Chi-square statistic, was < 0.001 (*p* < 0.05 corrected for multiple comparisons across 40 tracts). Patients are ordered based on the number of abnormal tracts. The color of the bars reflects the level of consciousness (LOC) at the day of MRI. MCS+: minimally conscious state plus; MCS-: minimally conscious state minus; PTCS: post-traumatic confusional state; VS: vegetative state. Patient IDs corresponding to those used in Table 1 (*P*2, …, *P*18) are indicated for each patient. **Right panel**: Boxplots showing the minimum, first quartile, median, third quartile and maximum number of significantly abnormal tracts for controls and patients. Outliers are defined as points greater than: third quartile + 1.5 * interquartile range. * significant difference (Wilcoxon rank sum test: *W* = 4.37, *p* = 1.22*e* − 05).

To investigate the accuracy of the multivariate pipeline at detecting TAI, we quantified its performance in classifying patients versus controls. We first computed the *D*_*M*_ for each control in a leave-one-out fashion and extracted the number of tracts that exceeded the critical value of the Chi-squared statistic at *α* = 0.001 for each control. On average 1.36 +/- 1.38 tracts appeared to have significantly abnormal *D*^2^_*M*_ values in controls (Figure 6, right panel). However, the mean *D*_*M*_ of these tracts was lower than the mean *D*_*M*_ of the abnormal tracts in controls (35 and 74 respectively). A Wilcoxon sum rank test showed a significant difference between the number of abnormal tracts (*W* = 4.37, *p* = 1.22*e* − 05) and between the *D*_*M*_ of these tracts (*W* = 7.11, *p* = 1.08*e* − 12) in patients versus controls. We then performed a ROC analysis to measure the performance of the pipeline in discriminating between patients and controls and found an accuracy of 0.91 (Supplementary figure S6). To visualize the difference in *D*^2^_*M*_ values between the two populations we plotted the probability density functions of *D*^2^_*M*_ for patients and controls. Figure 7 shows the probability density functions of *D*^2^_*M*_ values for the different sections of the corpus callosum for patients and controls. For subjects with severe TBI, *D*^2^_*M*_ values are shifted to the right suggesting a larger mean and greater degree of microstructural deviation from the normative reference group. Probability density functions of *D*^2^_*M*_ values for patients and controls for all 40 tracts are shown in supplementary figure S8.

**Figure 7.**
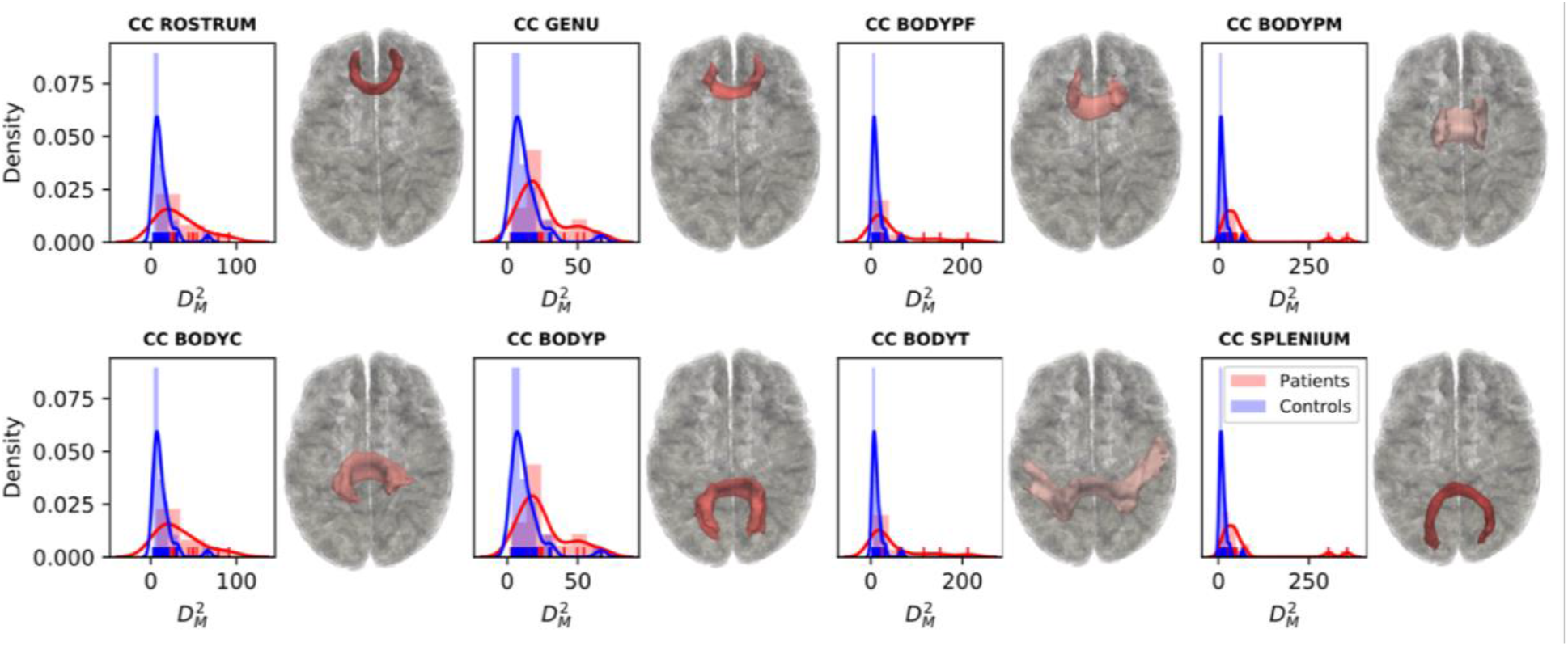
Density plots. Probability density functions of Mahalanobis distance (*D*^2^_*M*_) values are shown for patients (shown in red) and controls (shown in blue) for each of the different sections of the corpus callosum (CC). Representative 3D reconstructions of the correspondent WM tracts are shown to the right of the density plots. CC BODYC: central section of the body of the CC; CC BODYP: parietal section of the body of the CC; CC BODYPF: prefrontal section of the body of the CC; CC BODYPM: premotor section of the body of the CC; CC BODYT: temporal section of the body of the CC; CC GENU: genu of the CC; CC ROSTRUM: rostrum of the CC; CC SPLENIUM: splenium of the CC.

Finally, a Wilcoxon signed-rank test showed no significant difference in the number of abnormal tracts (*p* = 0.15) and in the mean *D*_*M*_ (*p* = 0.17) per patient when analyses were conducted including only the tracts that had been successfully reconstructed without the need for reinitialization (*i.e*., discarding tracts that were reinitialized) (Supplementary figure S8). The accuracy in discriminating between patients and controls also did not change (AUC = 0.91). This suggests that the low number of tracts that needed to be reinitialized across subjects did not significantly impact the overall individualized assessment of TAI.

### 3.4 Effects of head motion

The TMI and the four motion measures used to compute it are reported for each subject in supplementary Table S3. A nonparametric Wilcoxon sum rank test revealed a non-statistically significant difference in TMI between patients and controls (*W* = 1.91, *p* = 0.05). Results of the linear mixed effects model revealed that TMI did not significantly predict *D*^2^_*M*_ (*B* = −0.179, *SE* = 0.232, *t*(*df*) = −0.77(46.78), *p* = 0.44). However, group was a significant predictor of *D*^2^_*M*_ (*B* = 17.420, *SE* = 3.649, *t*(*df*) = 4.77(46.79) *p* < 0.001), with patients demonstrating significantly higher values than controls, as expected given how *D*^2^_*M*_ is calculated. See supplementary Table S4 for full results of the linear mixed effects model. Taken together, these results emphasize that the greater number of affected tracts detected for patients versus controls was not confounded by head motion in the scanner.

### 3.5 Relationship with behavioral measures of consciousness

Average *D*_*M*_ of affected tracts was not significantly correlated with GCS-Total, (*rho* = −0.16, *p* = 0.55), CRS-R Total (*rho* = −0.12, *p* = 0.65), days to command-following (*rho* = 0.32, *p* = 0.27), or days in coma (*rho* = 0.00, *p* = 1.00). Similarly, number of affected tracts was not significantly correlated with GCS-Total (*rho* = −0.29, *p* = 0.26), CRS-R Total (*rho* = −0.33, *p* = 0.19), days to command-following (*rho* = −0.13, *p* = 0.65), or days in coma (*rho* = 0.15, *p* = 0.56). Complete results from the non-parametric Spearman correlations are reported in supplementary table S5. The proposed linear regression models using average *D*_*M*_ of affected tracts and number of affected tracts to predict GCS-Total, CRS-R Total, days to command-following, and days in coma were not pursued, given that the correlation analyses did not reveal a significant association between these variables.

No significant relationships were found between LOC and average *D*_*M*_ of affected tracts (*B* = −0.02, *SE* = 0.01, *t* = −1.49, *p* = 0.136) or number of affected tracts (*B* = −0.029, *SE* = 0.052, *t* = −0.55, *p* = 0.582). The univariate ordinal logistic regressions testing the association between LOC and age (*B* = −0.020, *SE* = 0.047, *t* = −0.42, *p* = 0.67), TMI (*B* = 0.04, *SE* = 0.041, *t* = 0.97, *p* = 0.332), and days to MRI (*B* = 0.15, *SE* = 0.08, *t* = 1.79, *p* = 0.073) did not identify any confounding relationships, and thus, it was not necessary to include these variables as covariates in the model.

All statistical analyses were repeated using only the tracts that were successfully reconstructed without the need for reinitialization and results did not differ (Supplementary note, supplementary table S6).

## 4. Discussion

In this prospective observational study, we propose an automated pipeline that combines automated tractography reconstructions with multivariate analysis of along-tract diffusion metrics to assess the presence of TAI in individual patients with acute severe TBI. We show that i) TRACULA (Maffei et al., 2021; Yendiki et al., 2011) provides automated reconstruction of WM tracts from dMRI data in critically ill patients with acute severe TBI, even in the presence of large focal lesions; ii) the multivariate analysis detected at least one abnormal WM tract per patient and could accurately differentiate patients from healthy controls (AUC = 0.91). These proof-of-principle results suggest that TRACULA can be used by clinicians for acute detection of TAI burden in critically ill patients. This information can complement more conventional MRI exams, inform prognosis, guide therapeutic decision-making, and be incorporated into the standardized clinical assessment. We discuss the clinical implications and technical limitations of the current results and provide further directions for optimization and translation of the proposed pipeline.

With respect to TAI detection, the automated pipeline found that, on average, 9.07 +/- 7.91 tracts were abnormal in patients with TBI. The individualized profiles of WM damage highlighted the degree of heterogeneity in magnitude, number, and location of WM changes in patients (Figures 5,6 and supplementary figure S3). This finding builds on recent studies showing that diffusion measures can be used to identify the neuroanatomic distribution and severity of TAI at the individual level (Jolly et al., 2020), an important goal given the well-established heterogeneity of this condition (Douglas et al., 2019). Despite the heterogeneity of the specific WM tracts that identified as abnormal across patients, and the fact that no clear pattern of WM damage was visible within LOC groups (Figure 5), the multivariate individual analysis revealed commissural connections to be more significantly and more frequently affected compared to projection and association fibers across all LOC (Figure 5, supplementary figures S4, S5). Specifically, the premotor, parietal, and temporal sections of the CC were the most affected, consistent with previous histological and neuroimaging studies in humans with severe TBI (Moen et al., 2014; Nolan et al., 2020; Ubukata et al., 2016) and with studies in animal models of TAI (Baker et al., 2004; Gennarelli et al., 1982). Changes in diffusion measures in these WM tracts reflect a series of pathological events caused by shearing and straining forces exerted on the axons during TBI. Previous studies investigating the association between dMRI and histological measures reported correlations between decreased FA and axonal degeneration, between increased MD and axonal demyelination, but also between increased FA and neuroinflammation (Hutchinson et al., 2018). In this work we combined along-tract measures of both FA and MD, as multivariate approaches have shown higher power in distinguishing different clinical populations from controls (Dean et al., 2017). Therefore, changes in WM tracts in TBI patients might include both or either FA/MD increase or decrease and reflect a combination of degeneration, demyelination, or inflammatory mechanisms. Future studies are needed to investigate the prognostic value of different combinations of dMRI metrics.

Interestingly, individual multivariate measures of TAI did not show a significant relationship with behavioral measures of consciousness (*i.e*., GCS-Total, CRS-R Total, days to command-following, days in coma). There are several potential explanations for this observation. First, many clinical factors beyond TAI influence a patient’s LOC in the ICU, including other traumatic lesions (*e.g*., contusions and subdural hemorrhages) and subclinical seizures, as well as toxic, metabolic, endocrinologic or infectious causes of encephalopathy. Second, widespread tract disruption in the brainstem may have confounded any associations between the global burden of TAI and behavioral measures of consciousness. TRACULA does not yet provide automated reconstructions of brainstem tracts – a key direction for future work in this field, especially as the presence of TAI in brainstem pathways can cause altered consciousness (Edlow et al., 2013; Snider et al., 2019). Third, the small sample size and the variability in days to MRI across patients, together with the high number of tracts being tested, could have underpowered our analyses. Nevertheless, given that prior studies have identified correlations between acute tractography data and long-term cognitive function (Wang et al., 2008), these data provide the basis for future studies with larger sample sizes to test whether multivariate tract-specific measurements in the ICU predict recovery.

TRACULA provided automated reconstruction of 93% of the WM tracts, on average, without the need of manual intervention, offering potential for dissemination to a broad range of hospital settings without the need for local tractography expertise. However, the potential clinical translation of this pipeline for TAI detection in the ICU will depend upon additional methodological and logistical factors that require further study. While on average TRACULA could successfully reconstruct 37.5 of 40 tracts without the need of manual intervention, an average of 2.4 tracts resulted in failed or partial reconstructions requiring reinitialization in 12 patients (Supplementary table 1). Even if our results did not change when using only the WM tracts that did not require reinitialization, this step relied upon a visual quality assessment from a tractography expert that would limit TRACULA’s applicability to clinical settings. Furthermore, some reinitialized tracts could not be successfully reconstructed due to the presence of extensive focal lesions in two patients. While it has been previously shown that TRACULA is robust to errors in the anatomical segmentation (Zöllei et al., 2019), large lesions can lead to missing labels that can affect tract reconstructions, because prior probabilities are based on the relative positions of the bundles with respect to the labels. Further optimization of the TRACULA pipeline will include i) improved automated assessment of tract reconstructions and ii) correct handling of missing FreeSurfer anatomical labels, including in cases where the FreeSurfer “recon-all” pipeline fails due to the presence of lesions (*e.g*., Patient 1 in this study). Additionally, the dMRI data used in this study were acquired with high angular resolution (*b* = 2,000 *s*/*mm*^2^, 60 diffusion-encoding directions) and relatively high spatial resolution (2 mm isotropic) compared to more standard clinical diffusion sequences (Chilla et al., 2015), leading to a longer acquisition time (∼ 9 minutes). Thus, while obtained on a clinical scanner, the dMRI data collected in this study were likely of higher quality than dMRI data acquired on clinical scanners in most hospitals, and it will be important to determine whether automated reconstruction of WM tracts with TRACULA is robust in the setting of lower-resolution, lower-quality dMRI data in the future. It is also important to consider that the individualized assessment proposed here can only be performed comparing patients to a control cohort scanned with the same dMRI sequence. For this pipeline to be applied in a clinical setting that does not have data from healthy controls, future optimizations of this pipeline should explore its robustness to dMRI data acquired on a different scanner and with a different dMRI sequence. Furthermore, although there was no statistically significant difference in age between the patient and control cohorts, the controls were not enrolled with an even distribution of age and sex. Identification of optimal characteristics and sample sizes for control cohorts is an area of active inquiry (Jolly et al., 2020), and future studies may leverage large normative databases such as the Human Connectome Project in this effort (Bookheimer et al., 2019; Harms et al., 2018). Finally, it is important to consider that some patients with acute severe TBI are not stable enough to travel to an MRI scanner, and thus this pipeline may not be feasible to use until these patients reach the subacute stage of care.

In summary, this study demonstrates the feasibility and utility of an automated tractography pipeline for detecting TAI in the acute injury phase of severe TBI. Quantitative analysis of patient-specific TAI burden in the ICU could increase the accuracy of outcome prediction in this population and guide decisions regarding the provision of life-sustaining treatment and rehabilitation.

## Data Availability

The TRACULA version and the training dataset are included in FreeSurfer 7.2 (https://github.com/freesurfer/freesurfer/tree/fs-7.2). Extensive documentation and tutorials are available on the FreeSurfer wiki (https://surfer.nmr.mgh.harvard.edu/fswiki/Tracula). Part of the raw DWI data are available at https://openneuro.org/datasets/ds003367/versions/1.0.0. All remaining DWI data produced in the present study are available upon request to the authors.

## Acknowledgments

We thank the nursing staffs of the Massachusetts General Hospital Neurosciences ICU, Multidisciplinary ICU, and Surgical ICU. We also thank the Massachusetts General Hospital MRI technologists for assistance with data acquisition. We are grateful to the patients and families in this study for their participation and support.

## Disclaimer

The views expressed in this manuscript are entirely those of the authors and do not necessarily reflect the views, policy, or position of the United States Government, Department of Defense, or United States Special Operations Command.

