## Supplementary Information for "Automated detection of axonal damage along white matter tracts in acute severe traumatic brain injury"

**Supplementary Tables**

| ID | Number of TRACULA Reconstructed Tracts | | | | Focal Lesions on Acute Structural MRI |
|---|---|---|---|---|---|
| | i) Complete | i) Reinitialized | iii) Incomplete | iv) Failed | |
| P1 | 0* | N/A | N/A | N/A | R frontal and parietal contusions; small R frontal subdural hemorrhage |
| P2 | 40 | 0 | 0 | 0 | L frontal EVD tract |
| P3 | 40 | 0 | 0 | 0 | None |
| P4 | 38 | 2 [MCP, LH CST] | 2 [MCP, LH CST] | 0 | Bifrontal contusions; R frontoparietal contusion |
| P5 | 37 | 3 [LH CST, ACOMM, RH FAT] | 0 | 0 | L frontal contusion |
| P6 | 40 | 0 | 0 | 0 | L frontal contusion |
| P7 | 33 | 7 [CC BODYPM, ACOMM, LH SLF1, RH CST, RH FAT, LH FAT, RH SLF1, | 0 | 0 | None |
| P8 | 37 | 3 [LH CST, RH CST, RH MLF] | 0 | 0 | R posterior temporal contusion; L anterior temporal contusion |
| P9 | 37 | 3 [RH CST, RH ECM, RH SLF1] | 0 | 0 | Hemorrhage in splenium of corpus callosum |
| P10 | 37 | 3 [ACOMM, RH ECM. RH UF] | 0 | 2 [ACOMM, RH UF] | R anterior temporal contusion; L tentorial subdural hemorrhage |

| | | | | | |
|---|---|---|---|---|---|
| P11 | 38 | 2 [LH CBD, ACOMM] | 0 | 0 | L temporo-parietal contusion; R thalamic hemorrhage |
| P12 | 40 | 0 | 0 | 0 | R anterior temporal contusion |
| P13 | 37 | 3 [LH CST, RH CST, LH CBV] | 0 | 0 | L frontal contusion |
| P14 | 34 | 6 [RH SLF1, ACOMM, CC BODY PF, RH SLF2, RH FAT, RH UF] | 1 [RH SLF1] | 3 [ACOMM, CC BODY PF, RH SLF2] | R frontal contusion; L frontal and L mesial temporal contusions |
| P15 | 39 | 1 [ LH CBV] | 0 | 0 | L insular contusion; R frontal EVD tract |
| P16 | 40 | 0 | 0 | 0 | None |
| P17 | 35 | 5 [RH CBV, RH CBD, RH EMC, RH OR, RH FAT] | 0 | 0 | R anterior temporal and L posterior temporal contusions; R frontal and L temporal subdural hemorrhages. |
| P18 | 36 | 4 [CC BODYPM, CC ROSTRUM, LH FAT, LH CST] | 0 | 0 | L temporal contusion; L frontal EVD tract. |
| **Mean** | **37.52** | **2.47** | **0.17** | **0.29** | - |
| **SD** | **2.18** | **2.182821 2.18** | **0.5285954 0.52** | **0.848875 0.84** | - |

**Supplementary Table S1.** The table reports **i)** the number of tracts for each patient that were reconstructed completely and accurately in TRACULA without the need for reinitialization; **ii)** the tracts that were reinitialized upon visual inspection; **iii)** the tracts that resulted in partial reconstructions after reinitialization; **iv)** the tracts that resulted in failed reconstructions after reinitialization. The presence and location of focal lesions, along with the TBI mechanisms and LOC at MRI are also reported. *In this subject (P1) the FreeSurfer "recon-all" pipeline failed due to lesion and it was thus not possible to run the TRACULA pipeline. EVD = external ventricular drain. LOC: level of consciousness; MCS: minimally conscious state; PTCS post-traumatic confusional state; SD: standard deviation; VS: vegetative state. **Tract abbreviations**: ACOMM: anterior commissure; AF: arcuate fasciculus; AR: acoustic radiation; ATR: anterior thalamic radiation; CC BODYC: central section of the body of the CC; CC BODYP: parietal section of the body of the CC; CC BODYPF: prefrontal section of the body of the CC; CC BODYPM: premotor section of the body of the CC; CC BODYT: temporal section of the body of the CC ; CC GENU: genu of the CC; CC ROSTRUM: rostrum of the CC; CC SPLENIUM: splenium of the CC; CBD: dorsal portion of cingulum bundle; CBV: ventral portion of the cingulum bundle; CC: corpus callosum; CST: cortico-spinal tract; EMC: extreme capsule; FAT: frontal Aslant tract; ILF: inferior longitudinal fasciculus; LH: left hemisphere; ILF: middle longitudinal fasciculus; OR: optic radiation; RH: right hemisphere; SLF I,II,III: first, second, and third branch of the superior longitudinal fasciculus; UF: uncinate fasciculus.

| Metric | Tract | W | p-value |
|---|---|---|---|
| FA_0 | CC BODYPM | 0.8807721 | 0.0017522 |

| | | | |
|---|---|---|---|
| FA_0 | RH UF | 0.916552 | 0.0147221 |
| FA_0 | LH AR | 0.9212808 | 0.0198839 |
| FA_0 | LH EMC | 0.9320967 | 0.0401684 |
| FA_0 | RH SLF2 | 0.9262095 | 0.0273236 |
| FA_0 | RH CBV | 0.9236934 | 0.023218 |
| FA_0 | LH CBV | 0.9285724 | 0.031871 |
| FA_1 | RH SLF3 | 0.8820088 | 0.0018781 |
| FA_1 | CC GENU | 0.8516014 | 0.0003693 |
| FA_1 | RH CBD | 0.9334387 | 0.0438909 |
| FA_1 | CC BODYC | 0.9145991 | 0.0130204 |
| FA_1 | ACOMM | 0.9073055 | 0.0082862 |
| FA_1 | LH SLF2 | 0.9322933 | 0.0406927 |
| FA_2 | RH AR | 0.9312109 | 0.0378918 |
| FA_2 | RH CBD | 0.9313078 | 0.0381343 |
| FA_2 | CC SPLENIUM | 0.9324933 | 0.0412334 |
| FA_2 | RH SLF1 | 0.9267231 | 0.0282511 |
| FA_2 | RH CST | 0.8761603 | 0.0013563 |
| FA_2 | CC BODYC | 0.9220161 | 0.0208435 |
| FA_2 | ACOMM | 0.9217503 | 0.0204911 |
| FA_3 | RH ATR | 0.9114721 | 0.0107128 |
| FA_3 | MCP | 0.9108586 | 0.010313 |
| FA_3 | LH CBD | 0.8941281 | 0.0037642 |
| FA_3 | CC BODYC | 0.9137886 | 0.0123761 |
| FA_3 | LH MLF | 0.9151347 | 0.0134654 |
| FA_3 | RH CBV | 0.8393757 | 0.0002004 |
| FA_3 | LH CBV | 0.8521664 | 0.0003801 |
| MD_0 | LH SLF3 | 0.8694882 | 0.0009427 |
| MD_0 | CC BODYPF | 0.8447102 | 0.0002609 |
| MD_0 | LH FAT | 0.9088278 | 0.0090978 |
| MD_0 | RH SLF3 | 0.7399372 | 2.79E-06 |
| MD_0 | CC GENU | 0.928408 | 0.0315307 |
| MD_0 | LH AR | 0.9332277 | 0.0432828 |
| MD_0 | RH CBD | 0.8790717 | 0.0015936 |
| MD_0 | RH SLF2 | 0.9299652 | 0.0349143 |
| MD_0 | LH OR | 0.8745909 | 0.0012442 |
| MD_0 | LH CST | 0.928028 | 0.0307579 |
| MD_0 | RH CBV | 0.8323573 | 0.0001426 |
| MD_0 | CC BODYT | 0.8907428 | 0.0030908 |
| MD_0 | LH CBV | 0.8262106 | 0.0001064 |
| MD_1 | CC BODYPF | 0.9043596 | 0.006925 |
| MD_1 | RH MLF | 0.9168459 | 0.0149978 |
| MD_1 | RH UF | 0.903201 | 0.0064562 |
| MD_1 | LH ILF | 0.907908 | 0.0085979 |
| MD_1 | LH UF | 0.9124511 | 0.0113851 |
| MD_1 | ACOMM | 0.8992136 | 0.0050829 |
| MD_1 | RH AF | 0.8600684 | 0.0005717 |
| MD_1 | LH MLF | 0.8642138 | 0.0007111 |
| MD_1 | CC BODYT | 0.917487 | 0.0156181 |
| MD_2 | RH ATR | 0.9191426 | 0.0173478 |
| MD_2 | LH OR | 0.5514259 | 1E-08 |

| | | | |
|---|---|---|---|
| MD_2 | RH SLF1 | 0.9067864 | 0.0080273 |
| MD_2 | LH ILF | 0.7205661 | 1.36E-06 |
| MD_2 | RH CST | 0.9178838 | 0.0160154 |
| MD_2 | LH SLF2 | 0.9212015 | 0.0197832 |
| MD_2 | CC BODYT | 0.9194121 | 0.0176478 |
| MD_3 | LH SLF3 | 0.8728746 | 0.0011327 |
| MD_3 | RH SLF3 | 0.8530392 | 0.0003974 |
| MD_3 | RH FAT | 0.8866142 | 0.0024378 |
| MD_3 | RH ILF | 0.7975626 | 2.901E-05 |
| MD_3 | LH SLF1 | 0.9349226 | 0.0484249 |
| MD_3 | LH AR | 0.8107626 | 5.212E-05 |
| MD_3 | RH AR | 0.8252399 | 0.0001016 |
| MD_3 | RH EMC | 0.9247809 | 0.0249073 |
| MD_3 | LH OR | 0.7463136 | 3.56E-06 |
| MD_3 | LH ILF | 0.7221288 | 1.44E-06 |
| MD_3 | ACOMM | 0.9195157 | 0.0177646 |
| MD_3 | LH MLF | 0.8449381 | 0.0002639 |
| MD_3 | RH CBV | 0.8811756 | 0.0017923 |

**Supplementary Table S2.** The table reports the tracts that for each dMRI metric (FA_0, FA_1, FA_2, FA_3, MD_0, MD_1, MD_2, MD_3) showed that the distribution of their values across the controls departed significantly from normality (Shapiro-Wilk test, $p < 0.05$). W- and p-values are reported for each tract. For these tracts, a rank-based inverse normal transformation was applied to the data for both controls and patients. For a complete list of WM tract abbreviations see Supplementary table S1.

| Subject | Group | Average Rotation | Average Translation | Portion of Slices with signal drop-out | Drop-out Severity | TMI |
|---|---|---|---|---|---|---|
| 1 | Controls | 0.001727 | 0.418448 | 0.000000 | 1.00000 | 1.03797 |
| 2 | Controls | 0.001329 | 0.319263 | 0.000000 | 1.00000 | -0.197668 |
| 3 | Controls | 0.003071 | 0.355562 | 0.000000 | 1.00000 | 1.82382 |
| 4 | Controls | 0.001846 | 0.352028 | 0.000000 | 1.00000 | 0.589744 |
| 5 | Controls | 0.001936 | 0.452903 | 0.000000 | 1.00000 | 1.53774 |
| 6 | Controls | 0.001706 | 0.384086 | 0.000000 | 1.00000 | 0.724929 |
| 7 | Controls | 0.001317 | 0.949004 | 0.000000 | 1.00000 | 5.15599 |
| 8 | Controls | 0.001924 | 0.282322 | 0.000000 | 1.00000 | 0.0722552 |
| 9 | Controls | 0.001469 | 0.240727 | 0.000000 | 1.00000 | -0.7291 |
| 10 | Controls | 0.001356 | 0.428688 | 0.000000 | 1.00000 | 0.760612 |
| 11 | Controls | 0.002197 | 0.274383 | 0.000000 | 1.00000 | 0.272836 |
| 12 | Controls | 0.001266 | 0.269774 | 0.000000 | 1.00000 | -0.681865 |
| 13 | Controls | 0.001247 | 0.356736 | 0.000000 | 1.00000 | 0.0403459 |
| 14 | Controls | 0.001499 | 0.230597 | 0.000000 | 1.00000 | -0.894333 |
| 15 | Controls | 0.001676 | 0.230809 | 0.000000 | 1.00000 | -0.689606 |
| 16 | Controls | 0.001565 | 0.321352 | 0.000000 | 1.00000 | -0.0188977 |
| 17 | Controls | 0.002104 | 0.260174 | 0.000000 | 1.00000 | 0.0574637 |
| 18 | Controls | 0.001334 | 0.208548 | 0.000000 | 1.00000 | -1.13678 |
| 19 | Controls | 0.002046 | 0.329952 | 0.000000 | 1.00000 | 0.597815 |
| 20 | Controls | 0.002134 | 0.232391 | 0.000000 | 1.00000 | -0.146972 |
| 21 | Controls | 0.001854 | 0.273294 | 0.000000 | 1.00000 | -0.0734511 |
| 22 | Controls | 0.003250 | 0.242338 | 0.345349 | 1.15499 | 1.03516 |
| 23 | Controls | 0.002197 | 0.196277 | 0.000000 | 1.00000 | -0.392795 |
| 24 | Controls | 0.002040 | 0.234621 | 0.000000 | 1.00000 | -0.219742 |
| 25 | Controls | 0.001960 | 0.286823 | 0.000000 | 1.00000 | 0.146285 |
| 26 | Controls | 0.001193 | 0.243690 | 0.000000 | 1.00000 | -0.976021 |
| 27 | Controls | 0.001523 | 0.237298 | 0.000000 | 1.00000 | -0.705873 |
| 28 | Controls | 0.001477 | 0.361191 | 0.000000 | 1.00000 | 0.304468 |
| 29 | Controls | 0.001706 | 0.271702 | 0.000000 | 1.00000 | -0.232331 |
| 30 | Controls | 0.002420 | 0.290654 | 0.000000 | 1.00000 | 0.631097 |
| 31 | Controls | 0.001344 | 0.282981 | 0.000000 | 1.00000 | -0.492588 |

| | | | | | | |
|---|---|---|---|---|---|---|
| 32 | Controls | 0.001116 | 0.237358 | 0.000000 | 1.00000 | -1.10542 |
| 33 | Controls | 0.001482 | 0.329762 | 0.213220 | 1.16272 | 0.0416457 |
| P2 | Patients | 0.002398 | 0.319144 | 0.000000 | 1.00000 | 0.851761 |
| P3 | Patients | 0.002449 | 0.242368 | 0.000000 | 1.00000 | 0.248332 |
| P4 | Patients | 0.014575 | 0.896906 | 0.134108 | 1.14880 | 17.7473 |
| P5 | Patients | 0.024903 | 1.010200 | 0.000000 | 1.00000 | 28.8665 |
| P6 | Patients | 0.002177 | 0.180212 | 0.000000 | 1.00000 | -0.549083 |
| P7 | Patients | 0.024673 | 0.908842 | 0.128260 | 1.07433 | 27.7767 |
| P8 | Patients | 0.001907 | 0.290166 | 0.000000 | 1.00000 | 0.121952 |
| P9 | Patients | 0.009026 | 0.282405 | 0.000000 | 1.00000 | 7.05599 |
| P10 | Patients | 0.008497 | 0.471386 | 0.000000 | 1.00000 | 8.14538 |
| P11 | Patients | 0.001599 | 0.275717 | 0.021505 | 1.00446 | -0.303609 |
| P12 | Patients | 0.000872 | 0.219660 | 0.000000 | 1.00000 | -1.49567 |
| P13 | Patients | 0.002152 | 0.269136 | 0.000000 | 1.00000 | 0.184329 |
| P14 | Patients | 0.001000 | 0.227638 | 0.000000 | 1.00000 | -1.30255 |
| P15 | Patients | 0.000921 | 0.215866 | 0.000000 | 1.00000 | -1.48044 |
| P16 | Patients | 0.007765 | 0.325994 | 0.000000 | 1.00000 | 6.1873 |
| P17 | Patients | 0.004763 | 0.348514 | 0.000000 | 1.00000 | 3.42733 |
| P18 | Patients | 0.013854 | 0.933081 | 0.000000 | 1.00000 | 17.3465 |

**Supplementary Table S3.** The table reports the MRI head motion measures as extracted from *eddy* in FSL 6.0.1. for each subject in the study. The total motion index (TMI) is a composite score of four motion measures: i) average volume-by-volume rotation, ii) average volume-by-volume translation, iii) percentage of slices with signal drop-out, and iv) signal drop-out severity.

| **Predictors** | **Estimates** | **Standard Error** | **t*(df)*** | ***p*** |
|---|---|---|---|---|
| (Intercept) | 9.37 | 3.79 | 2.47*(630)* | 0.013 |
| TMI | -0.18 | 0.23 | -0.77*(46.78)* | 0.445 |
| Group | 17.42 | 3.65 | 4.77*(46.79)* | <0.001 |
| CC BODYC | 8.19 | 4.67 | *1.75(1903)* | 0.079 |
| CC BODYP | 11.89 | 4.67 | 2.54*(1903)* | 0.011 |
| CC BODYPF | 10.46 | 4.70 | 2.23*(1903)* | 0.026 |
| CC CBODYPM | 16.85 | 4.67 | *3.60(1903)* | <0.001 |
| CC BODYT | 4.51 | 4.67 | 0.97*(1903)* | 0.334 |
| CC GENU | 1.50 | 4.67 | *0.32(1903)* | 0.748 |
| CC ROSTRUM | 3.96 | 4.67 | 0.85*(1903)* | 0.397 |
| CC SPLENIUM | 5.95 | 4.67 | 1.27*(1903)* | 0.203 |
| LH AF | 3.86 | 4.67 | 0.83(*1903)* | 0.409 |
| LH AR | 1.37 | 4.67 | 0.29(*1903)* | 0.768 |
| LH ATR | 2.68 | 4.67 | 0.57*(1903)* | 0.566 |
| LH CBD | 0.94 | 4.67 | 0.20*(1903)* | 0.840 |
| LH CBV | -2.42 | 4.67 | -0.52(*1903)* | 0.603 |
| LH CST | 6.31 | 4.67 | 1.34*(1903)* | 0.179 |
| LH EMC | 0.92 | 4.67 | 0.20(*1903)* | 0.844 |
| LH FAT | 1.88 | 4.67 | 0.40*(1903)* | 0.687 |

| | | | | |
|---|---|---|---|---|
| LH ILF | 1.52 | 4.67 | 0.32*(1903)* | 0.745 |
| LH MLF | -0.52 | 4.67 | 2.54*(1903)* | 0.911 |
| LH OR | -1.59 | 4.67 | -0.34*(1903)* | 0.733 |
| LH SLF1 | -1.52 | 4.67 | -0.33*(1903)* | 0.744 |
| LH SLF2 | -0.35 | 4.67 | -0.08*(1903)* | 0.940 |
| LH SLF3 | 2.10 | 4.67 | 0.45*(1903)* | 0.652 |
| LH UF | -0.92 | 4.67 | -0.20*(1903)* | 0.844 |
| MCP | 0.09 | 4.67 | 0.02*(1903)* | 0.985 |
| RH AF | -0.48 | 4.67 | -0.10*(1903)* | 0.918 |
| RH AR | 1.12 | 4.67 | 0.24*(1903)* | 0.810 |
| RH ATR | -0.19 | 4.67 | -0.04*(1903)* | 0.967 |
| RH CBD | 4.18 | 4.67 | 0.90*(1903)* | 0.370 |
| RH CBV | -2.94 | 4.67 | -0.63*(1903)* | 0.529 |
| RH CST | 1.63 | 4.67 | 0.35*(1903)* | 0.727 |
| RH EMC | 7.78 | 4.67 | 1.66*(1903)* | 0.096 |
| RH FAT | 10.52 | 4.67 | 2.25*(1903)* | 0.024 |
| RH ILF | 1.92 | 4.67 | 0.41*(1903)* | 0.681 |
| RH MLF | -1.97 | 4.67 | -0.42*(1903)* | 0.673 |
| RH OR | -3.51 | 4.67 | -0.75*(1903)* | 0.452 |
| RH SLF1 | -0.48 | 4.67 | -0.10*(1903)* | 0.917 |
| RH SLF2 | -0.25 | 4.67 | -0.05*(1903)* | 0.957 |

| | | | | |
|---|---|---|---|---|
| RH SLF3 | -1.08 | 4.67 | -0.23*(1903)* | 0.817 |
| RH UF | 0.92 | 4.67 | 0.20*(1903)* | 0.844 |

**Supplementary Table S4.** The table reports the parameter estimates, standard error, t-values, degrees of freedom, and p-values for the linear mixed effects regression model testing the relationship between TMI and D2M, while accounting for group (i.e., patient, control) and tract (i.e., 40 reconstructed tracts) as categorical variables, and including participant as a random intercept. Controls served as the reference level for the group variable and the anterior commissure served as the reference level for the tract variable. For a complete list of WM tract abbreviations see Supplementary table S1.

| Variable Type | Variable | GCS Total | CRS-R Total | Days to Command Following | Days in Coma | Average $D_M$ of affected tracts | Number of affected tracts | Age | TMI |
|---|---|---|---|---|---|---|---|---|---|
| Dependent Variables | GCS-Total | | | | | | | | |
| | CRS-R Total | **0.97** ***p* < 0.001** | | | | | | | |
| | Days to command -following | -0.49 *p = 0.073* | **-0.57** ***p* = 0.032** | | | | | | |
| | Days in coma | **-0.81** ***p* < 0.001** | **-0.82** ***p* < 0.001** | **0.58** ***p* = 0.029** | | | | | |
| Independent Variables | Average $D_M$ of affected tracts | -0.16 *p* = 0.547 | -0.12 *p* = 0.650 | 0.32 *p* = 0.267 | -0.001 *p* = 0.996 | | | | |
| | Number of affected tracts | -0.29 *p* = 0.260 | -0.33 *p* = 0.191 | 0.13 *p* = 0.651 | 0.151 *p* = 0.563 | 0.26 *p* = 0.321 | | | |
| Potential Confounding Variables | Age | -0.09 *p* = 0.721 | -0.12 *p* = 0.653 | 0.22 *p* = 0.456 | 0.33 *p* = 0.193 | -0.09 *p* = 0.729 | 0.12 *p* = 0.656 | | |
| | TMI | 0.44 *p* = 0.075 | 0.37 *p* = 0.142 | -0.27 *p* = 0.355 | -0.37 *p* = 0.140 | 0.03 *p* = 0.898 | 0.163 *p* = 0.533 | 0.03 *p* = 0.903 | |
| | Days to MRI | **0.54** ***p* = 0.026** | 0.42 *p* = 0.091 | -0.10 *p* = 0.740 | -0.22 *p* = 0.387 | -0.03 *p* = 0.903 | 0.31 *p* = 0.222 | 0.12 *p* = 0.649 | **0.53** ***p* = 0.028** |

**Supplementary Table S5**. Spearman correlation results using all the tracts – successfully reconstructed and reinitialized. The *rho* coefficient and *p-value* are reported for each pair of variables. Significant correlations at the *p* < 0.05 level are bolded. Note, three participants were missing data for days to command-following, and thus, those correlations were conducted with n = 14. All other correlations had n = 17. CRS-R = Coma Recovery Scale-Revised, GCS = Glasgow Coma Scale, TMI = Total Motion Index.

| Variable Type | Variable | GCS Total | CRS-R Total | Days to Command Following | Days in Coma | Average $D_M$ of affected tracts | Number of affected tracts | Age | TMI |
|---|---|---|---|---|---|---|---|---|---|
| Dependent Variables | GCS-Total | 1.00 | | | | | | | |
| | CRS-R Total | **0.97**<br>***p* < 0.001** | | | | | | | |
| | Days to command-following | -0.49<br>*p* = 0.073 | **-0.57**<br>***p* = 0.032** | | | | | | |
| | Days in coma | **-0.81**<br>***p* < 0.001** | **-0.82**<br>***p* < 0.001** | **0.58**<br>***p* = 0.029** | | | | | |
| Independent Variables | Average $D_M$ of affected tracts | -0.17<br>*p* = 0.508 | -0.14<br>*p* = 0.603 | 0.31<br>*p* = 0.281 | -0.001<br>*p* = 0.995 | | | | |
| | Number of affected tracts | -0.28<br>*p* = 0.270 | -0.32<br>*p* = 0.206 | 0.11<br>*p* = 0.716 | 0.15<br>*p* = 0.563 | 0.27<br>*p* = 0.302 | | | |
| Potential Confounding Variables | Age | -0.09<br>*p* = 0.720 | -0.12<br>*p* = 0.653 | 0.22<br>*p* = 0.456 | 0.33<br>*p* = 0.193 | -0.07<br>*p* = 0.778 | 0.09<br>*p* = 0.718 | | |
| | TMI | 0.44<br>*p* = 0.074 | 0.37<br>*p* = 0.141 | -0.27<br>*p* = 0.355 | -0.37<br>*p* = 0.140 | 0.05<br>*p* = 0.846 | 0.16<br>*p* = 0.530 | 0.03<br>*p* = 0.903 | |
| | Days to MRI | **0.54**<br>***p* = 0.025** | 0.42<br>*p* = 0.090 | -0.10<br>*p* = 0.740 | -0.22<br>*p* = 0.387 | -0.04<br>*p* = 0.873 | 0.32<br>*p* = 0.204 | 0.12<br>*p* = 0.649 | **0.53**<br>***p* = 0.028** |

**Supplementary Table S6**. Spearman correlation results using only the tracts that were successfully reconstructed in TRACULA without the need of reinitialization**.** The *rho* coefficient and *p-value* are reported for each pair of variables. Significant correlations at the $p < 0.05$ level are bolded. Three participants were missing data for days to command-following, and thus, those correlations were conducted with n = 14. All other correlations had n = 17. CRS-R = Coma Recovery Scale-Revised, GCS = Glasgow Coma Scale, TMI = Total Motion Index.

### Supplementary Note

Supplementary table S6 reports the correlation results using only the tracts that were successfully reconstructed in TRACULA without the need of reinitialization. These results do not differ in their interpretation from those obtained when also using both the tracts that were reconstructed in TRACULA without the need of reinitialization and the tracts that needed reinitialization (reported in the main text under *section 3.5)*.

**Supplementary Figures**

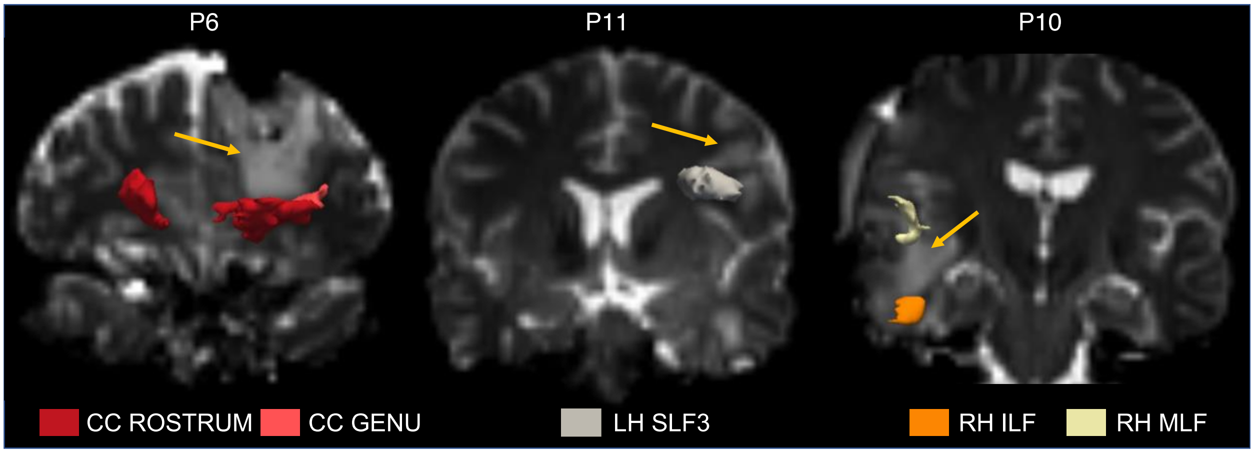


**Supplementary Figure S1.** Examples of successful TRACULA reconstructions in three patients with focal lesions. 3D tract reconstructions are shown on the b = 0 s/mm$^2$ dMRI volume thresholded at 20% of the maximum. Orange arrows point to the lesion. Coronal images are shown in radiological convention. Patient IDs correspond to table S2. CC GENU: genu of the corpus callosum; CC ROSTRUM: rostrum of the corpus callosum; LH SLF 3: third (most ventral) branch of the left superior longitudinal fasciculus. RH ILF: right inferior longitudinal fasciculus; RH MLF: right middle longitudinal fasciculus.

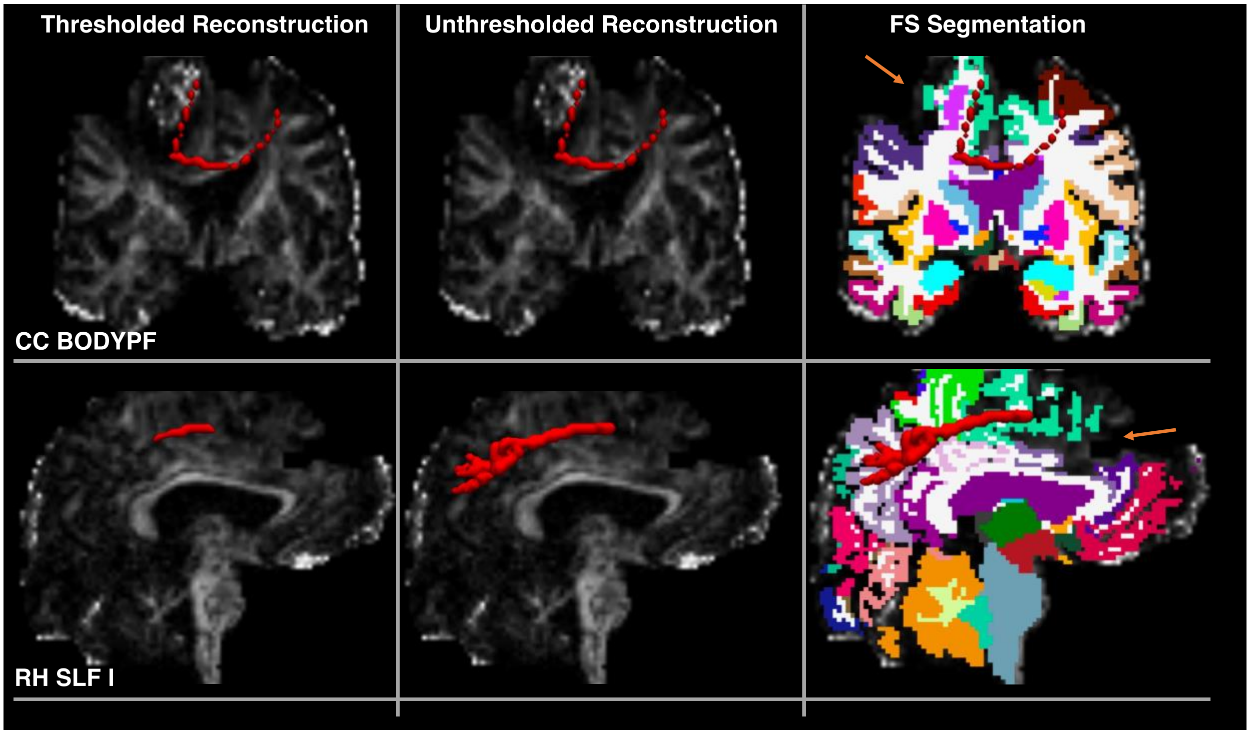


**Supplementary Figure S2.** Example of one failed and one partial TRACULA reconstructions for P14 (See table S2). RH SLFI: first (most dorsal) branch of the right superior longitudinal fasciculus. CC BODYPF: pre-frontal section of the corpus callosum. 3D tract reconstructions are shown on the fractional anisotropy scalar map for both thresholded (tract visitation maps thresholded at 20% of the maximum) and unthresholded posterior distributions. Unthresholded 3D reconstructions are also shown overlaid onto the FreeSurfer anatomical segmentation. Orange arrows point to regions where the segmentation was abnormal due to the presence of a large frontal focal lesion.

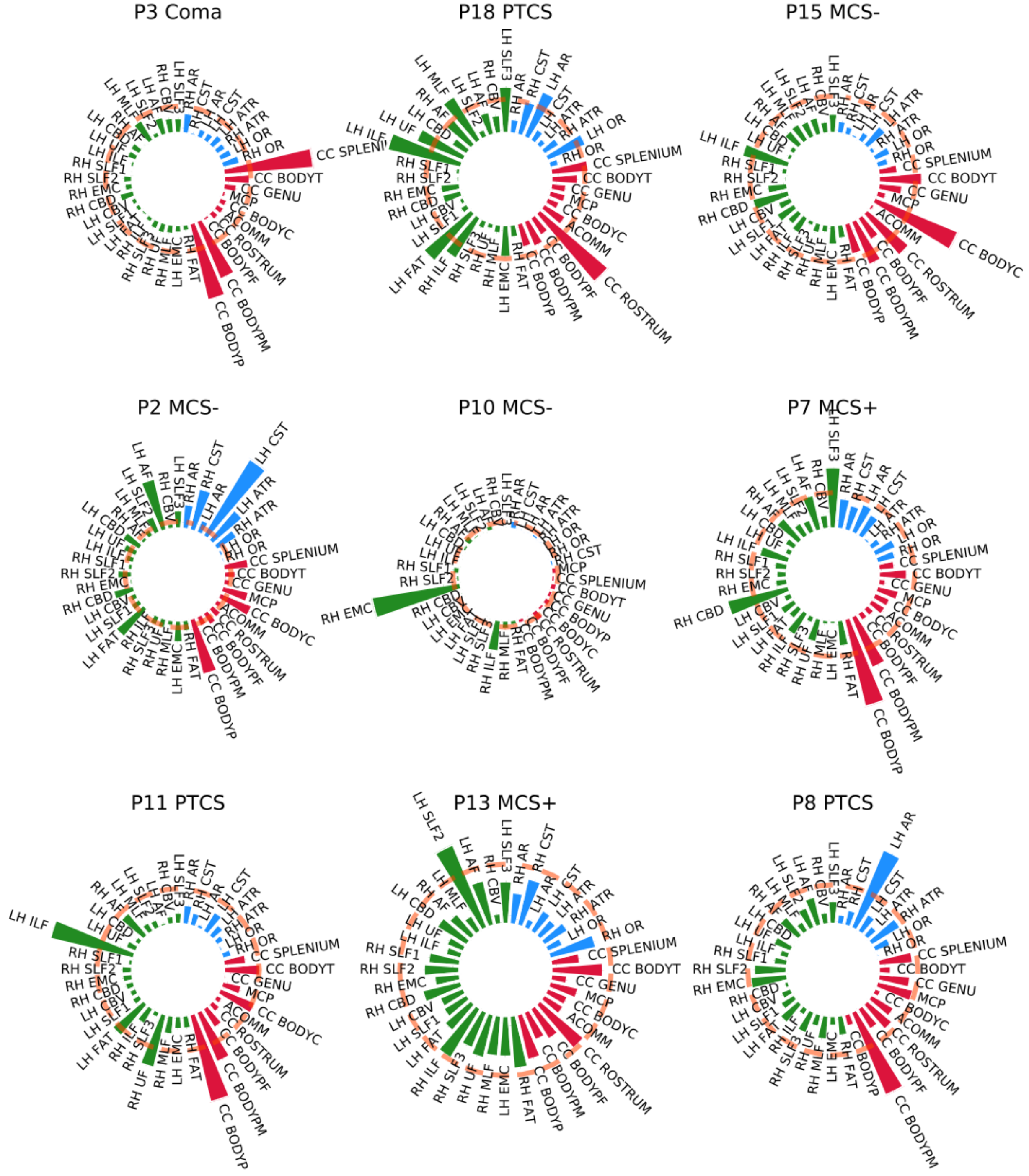
P3 Coma
P18 PTCS
P15 MCS-
P2 MCS-
P10 MCS-
P7 MCS+
P11 PTCS
P13 MCS+
P8 PTCS

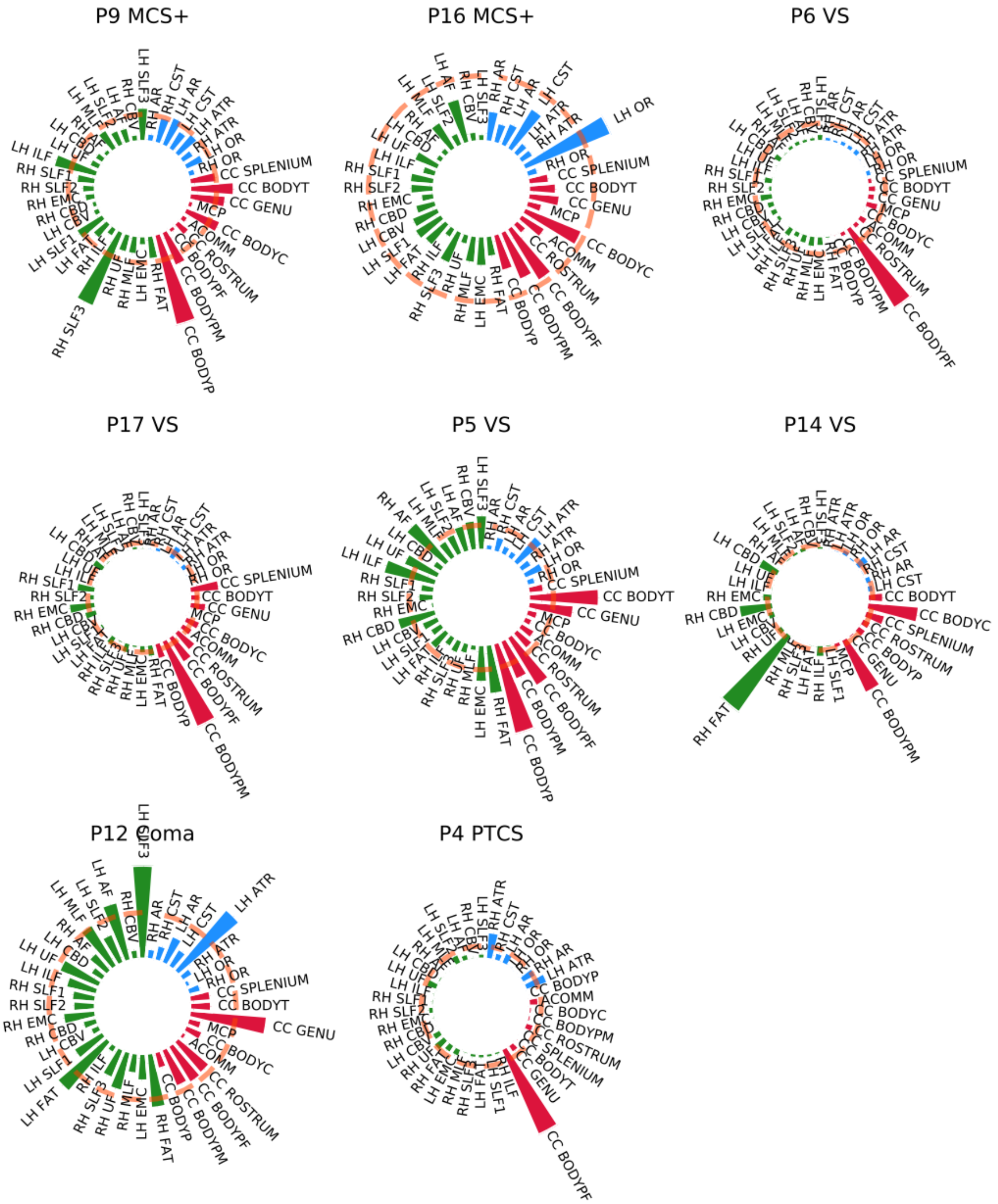


**Supplementary Figure S3. Individual Patient Profiles.** The figure shows the individual profiles for the 17 patients. The Mahalanobis distance for the 40 reconstructed tracts is plotted using polar bar plots. Tracts that resulted in either partial or failed reconstruction after reinitialization are not included in the individual profile of that patient. The pink dotted line corresponds to the critical value of the Chi-squared statistic at $\alpha = 0.001$. White matter tracts are grouped in commissural (red), associative (green), and projection pathways (blue). Tracts that resulted in either partial or failed reconstruction after reinitialization are not included. Level of consciousness at MRI for each patient is also reported (Coma; MCS+: minimally conscious state plus, evidence of language function (*e.g.,* reproducible movement to command, object recognition, intelligible verbalization); MCS-: minimally conscious state minus, no evidence of language function, but shows at least one behavior consistent with a minimally conscious state (*e.g.,*

automatic motor response); PTCS: post-traumatic confusional state; VS: vegetative state). For a complete list of WM tract abbreviations see table S2. Patient IDs correspond to table S2.

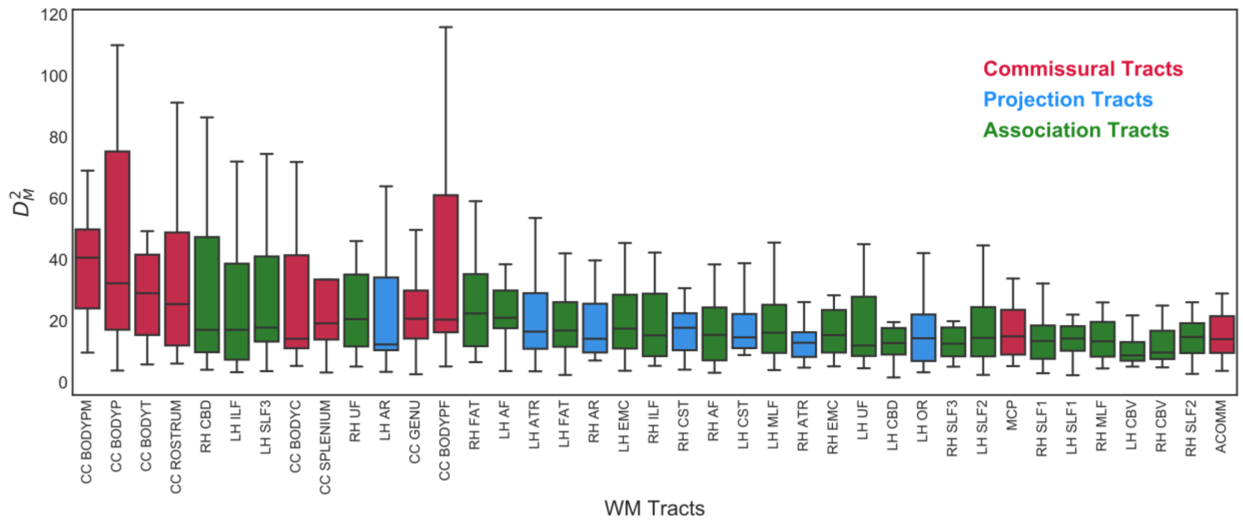


**Supplementary Figure S4.** The box plot reports the Mahalanobis distance ($D^2{}_M$) for each tract across all subjects with TBI. Bar colors indicate the tract group: commissural (red), projection (blue), association (green). Tracts that resulted in either partial or failed reconstruction after reinitialization are not included. For a complete list of white matter (WM) tract abbreviations see table S2.

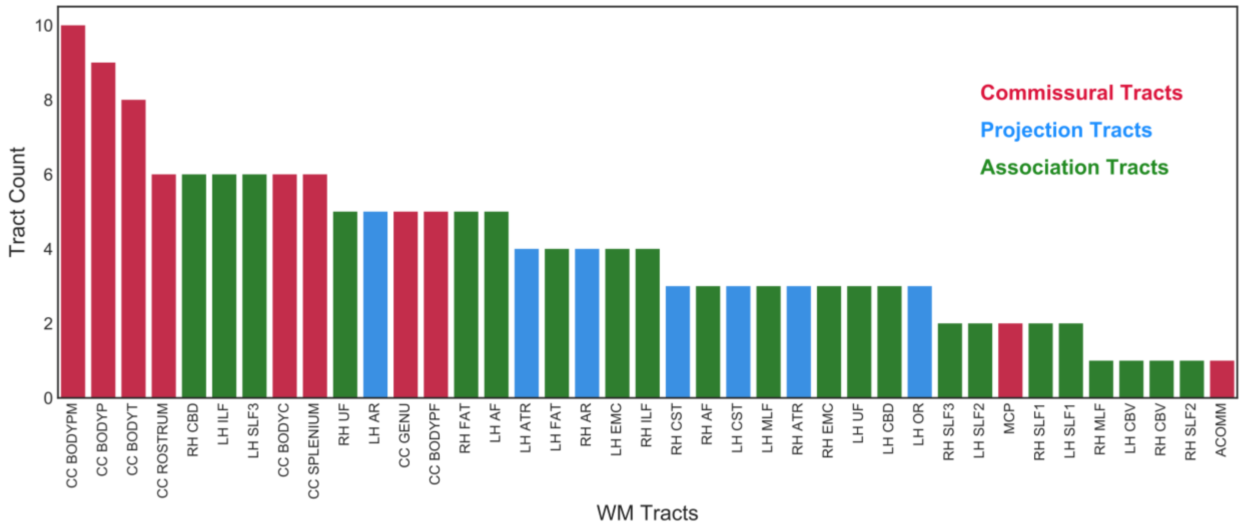


**Supplementary Figure S5.** The bar plot reports the number of times each tract resulted damaged across patients. Bar colors indicate the tract group: commissural (red), projection (blue), association (green). Tracts that resulted in either partial or failed reconstruction after reinitialization are not included. For a complete list of white matter (WM) tract abbreviations see table S2.

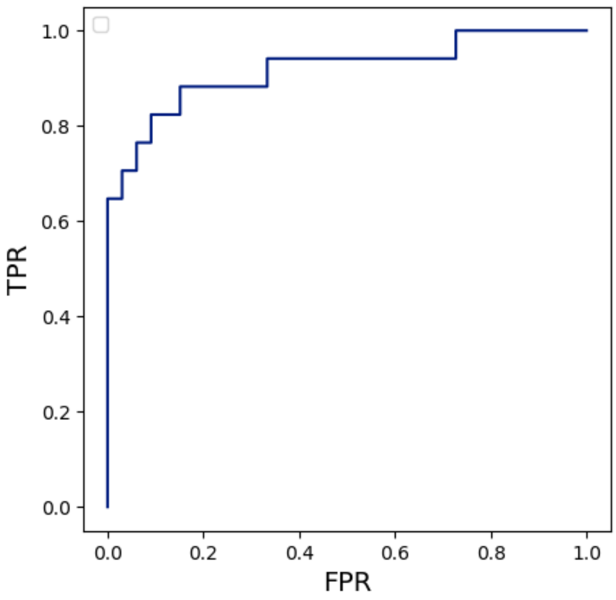


**Supplementary Figure S6.** Receiver operator characteristic (ROC) curve shows the performance of the multivariate pipeline in discriminating between patients and controls. FPR: false positive rate; TPR: true positive rate.

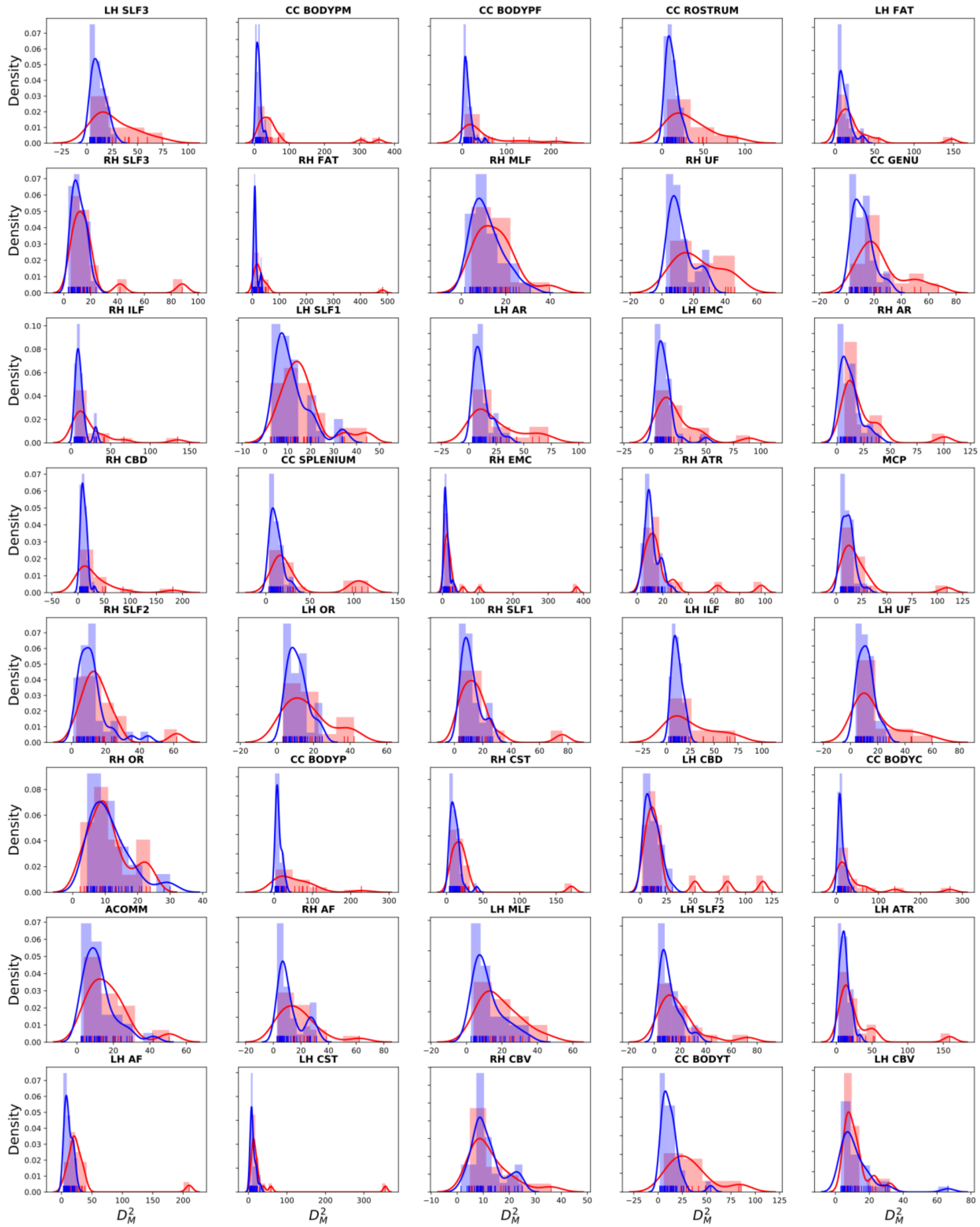


**Supplementary Figure S7.** Probability density functions of Mahalanobis distance values ($D^2{}_M$) are shown for patients (red) and controls (blue) for each of the 40 reconstructed tracts. For a complete list of WM tract abbreviations see table S2.

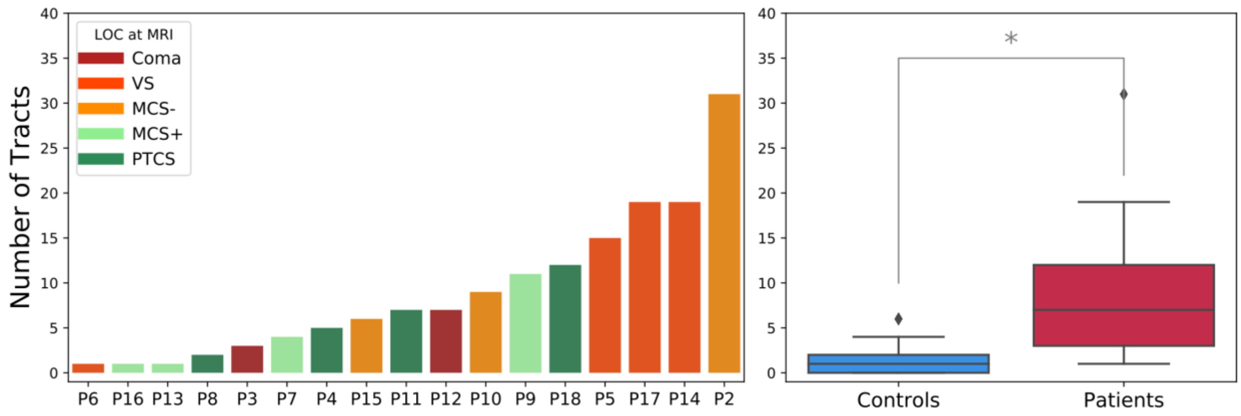


**Supplementary Figure S8.** The figure reports the results of the multivariate analysis when removing for each subject the tracts that needed to be reinitialized (i.e., using only the tracts that were successfully reconstructed in TRACULA without manual intervention) (See table S2). **Left panel)** The number of extreme WM tracts is shown for each subject with TBI. A tract was considered extreme if its $p - value$, correspondent to a Chi-square statistic, was $< 0.001$ (p < 0.05 corrected for multiple comparisons across 40 tracts). Patients are ordered based on the number of damaged tracts. The color of the bars reflects the level of consciousness (LOC) at the day of MRI. IDs corresponding to those used in Table S2 ($P2, \dots, P18$) are indicated for each patient. **Right panel)** Boxplots showing the number of extreme tracts for controls and patients. Asterisk indicate a significant difference (Wilcoxon rank sum test : $W = 4.36$, $p = 1.28e - 05$).
